# Using human genetic variation to estimate the effect of lipoprotein(a) lowering on pregnancy outcomes

**DOI:** 10.64898/2026.05.18.26351595

**Authors:** Helena Urquijo, Allison B Goldfine, Juan P Casas, Huilei Xu, Yoav Timsit, Mike Mendelson, Carolina Hache, Ieuan Jones, Daria Arustamian, Maria C Magnus, Tom R Gaunt, Deborah A Lawlor, Maria Carolina Borges

## Abstract

**Background:** Lipoprotein(a) (Lp[a]) is a genetically determined causal and independent cardiovascular risk factor and Lp(a) targeted therapies are being developed. However, evidence on the safety of substantial Lp(a) lowering during pregnancy is limited. We evaluated the impact of Lp(a) lowering on adverse pregnancy and perinatal outcomes (APPOs) using human genetic evidence.

**Material and Method:** We applied a drug-target Mendelian randomization (MR) approach using genetic variants associated with Lp(a) in the UK Biobank at the *LPA* locus to proxy pharmacological Lp(a) lowering. Summary-level APPO data were obtained from the MR-PREG collaboration, comprising up to 714,899 women across multiple studies. Twenty APPOs were included. Sensitivity analyses included adjustment for fetal genotype, alternative Lp(a) datasets, leave-one-study-out analyses, and exploration of Lp(a) genetic scores and individuals homozygous for *LPA* loss-of-function variants in the UK Biobank.

**Results:** Across 20 APPOs, MR estimates showed no strong evidence of causal effects, with no associations surviving false discovery rate *P*-value correction. Most estimates were close to null, including gestational hypertension, gestational diabetes, preeclampsia, miscarriage and neonatal intensive care unit admission. Some associations were slightly larger in magnitude but with wide confidence intervals: gestational age (mean difference 0.04 weeks, 95% CI 0.02–0.06 per 210nmol/L reduction in Lp[a]) and congenital malformation (OR 0.82, 95% CI: 0.72–0.94) in the protective direction of effect, and higher odds of stillbirth (OR 1.09, 95% CI: 1.00– 1.19) and low Apgar at 1 minute (OR 1.11, 95% CI: 0.99–1.24). Sensitivity analyses consistently supported the primary findings, with no evidence of increased maternal nor offspring risk in analyses adjusting for maternal– fetal genotype, across alternative exposure datasets, or in leave-one-study-out tests. Individual-level analyses of Lp(a) genetic score and *LPA* loss-of-function variants showed no associations, although power was limited.

**Conclusion:** These findings suggest that substantial lowering of Lp(a) is unlikely to increase APPO risk, although modest effects, particularly for rare outcomes, cannot be excluded.

## Introduction

Human epidemiologic and genetic evidence, particularly Mendelian Randomization (MR) studies, establish elevated plasma lipoprotein(a) (Lp[a]) as a genetically determined, independent, and putative causal risk factor for atherosclerotic cardiovascular disease (ASCVD), as well as calcific aortic valve stenosis^1^. Plasma Lp(a) levels are primarily genetically determined^2^ and are only minimally responsive to lifestyle interventions^3^. Existing therapeutic options are limited, with lipoprotein apheresis being moderately effective but impractical for patients requiring lifelong therapy, and conventional lipid-lowering agents such as proprotein convertase subtilisin/kexin type 9 (PCSK9) inhibitors and niacin achieving only modest reductions^4^. Consequently, there remains a clear unmet need for targeted therapies capable of achieving more substantial and sustained Lp(a) lowering.

Novel small nucleic acid therapies, including antisense oligonucleotide (ASO)-based and small interfering RNA (siRNA) agents targeting *LPA* mRNA expression in the liver, represent promising therapeutic approaches for reducing Lp(a) concentrations. Several phase III randomized controlled trials (RCTs) are currently underway in non-pregnant populations (ClinicalTrials.gov ID: NCT04023552, NCT05581303, NCT06292013). These candidate therapies have so far been shown to reduce mean Lp(a) levels by 80-95%, thus aiming to address the persistent cardiovascular risk associated with elevated Lp(a) levels despite contemporary risk management strategies^5–8^.

Because Lp(a) levels are largely genetically determined, future Lp(a)-lowering therapies may be used in relatively young individuals, making it important to understand any potential effects on human reproduction and development. This is increasingly relevant as more women enter pregnancy with established cardiovascular disease and use ASCVD medications around conception^9,10^. Individuals seeking to conceive, their partners, and clinicians need evidence to balance the risk of adverse pregnancy and perinatal outcomes (APPOs) against the potential increase in ASCVD risk from discontinuing treatment. However, conventional nonclinical safety studies are challenging because *LPA* is restricted to humans, apes, and Old-World monkeys, limiting the relevance of rodent models and complicating interpretation of non-human primate safety studies due to structural differences in the *LPA* gene with humans^11,12^. In light of these constraints, human genetic studies provide a powerful alternative to improve the current evidence base on the safety of this intervention during pregnancy.

MR is an instrumental variable (IV) analysis that leverages the random allocation of genetic variants at conception to investigate the effects of modifiable risk factors on health outcomes. This method mitigates confounding from socioeconomic, behavioural, or health-related factors that frequently bias traditional observational analyses^13^. The rationale of MR parallels that of RCTs: in RCTs, randomization determines treatment allocation and its effect on the outcome, assuming no alternative pathways. In MR, genetic variants act as a naturally occurring form of randomization, influencing the exposure and enabling causal inference under comparable assumptions.

Given the predominantly genetic basis of inter-individual variation in Lp(a) concentrations, in this study we exploit genetics to proxy Lp(a) levels, including loss-of-function (LoF) variants in *LPA*, to assess whether low Lp(a) levels have a causal role in APPO risk. These insights may help inform on the potential effects in pregnancy that may or may not be observed with pharmacologic Lp(a) lowering interventions for atherosclerotic risk reduction.

## Methods

### Study design

This study employs a two-sample MR design to evaluate the causal effect of lowering Lp(a) levels on 20 APPOs. Of these, 19 APPOs were selected out of the 35 available outcomes from the Mendelian Randomization in Pregnancy (MR-PREG) collaboration on the basis of being (i) key safety outcomes or (ii) cardiometabolic traits that could plausibly be affected by Lp(a) levels. Additionally, we included postpartum haemorrhage (PPH; not an outcome in MR-PREG), from a publicly available GWAS, due to homology between *LPA* and plasminogen (*PLG)*, and the established role of *PLG* in fibrinolytic pathways^14^. Genetic variants within the *LPA* locus, strongly associated with serum Lp(a) levels, are used as instrumental variables to mimic pharmacological modulation of Lp(a).

The primary analysis uses the generalized inverse-variance weighted MR method, leveraging summary statistics from large-scale genome-wide association studies (GWAS) of Lp(a) and APPOs. Sensitivity analyses were planned from the outset to investigate key sources of potential biases, such as due to outlying studies, sample overlap between exposure and outcome datasets, and confounding by linkage disequilibrium or fetal inheritance of maternal genotype. Analyses were repeated using coronary artery disease as a positive control outcome for comparison.

Additional exploratory analyses were prespecified and undertaken using individual-level UK Biobank (UKB) data, including thrombotic events and genetically predicted extremely low Lp(a) levels (equivalent of plausible levels resulting from Lp(a)-lowering therapies currently under clinical testing).

All features of the design, including exploratory analyses, were documented and justified before analyses began. The study design and data flow is illustrated in **Figure 1**.

**Figure 1:**
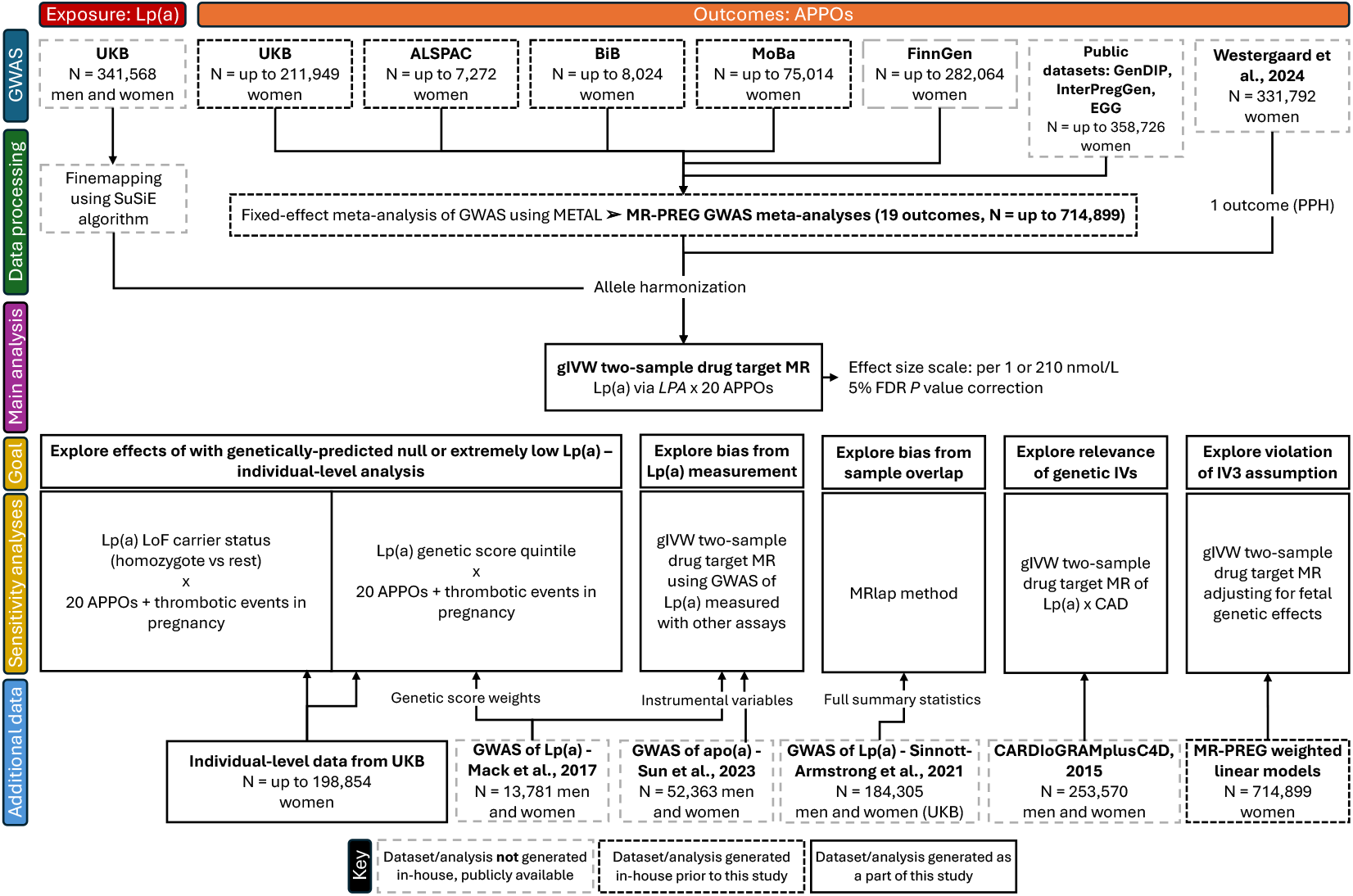
Diagram of study design and data sources.

### Data sources

#### Instrumental variables

The *LPA* gene encodes the apolipoprotein(a) that covalently binds to a cholesterol-rich low-density lipoprotein (LDL)-like particle to form Lp(a). For genetic instruments, we used 29 finemapped SNPs in the *LPA* region, previously identified as having the highest posterior inclusion probability (PIP) using SuSiE, a Bayesian finemapping method^15^. These 29 SNPs, combined in a genetic score, explained 68% of the phenotypic variance of Lp(a) concentration in a sample of 341,568 UK Biobank participants^16^. The 29 genetic variants finemapped by Shi et al. (2024) are listed in **Table S1**. Some SNPs were not present in MR-PREG data due to low frequency (MAF < 1%) or failing quality control (QC). Where possible, we used tag variants in high linkage disequilibrium (LD) (r^2^ > 0.8). The proportion of outcomes for which we had effect estimates for the original SNP or a tag variant are also shown in **Table S1**. The variation in Lp(a) explained by the IVs ranged from r^2^ ≈ 0.41 to 0.63, according to the outcome. The final number of SNPs included in the IV, along with the variance explained and F-statistic, are reported with each MR estimate.

#### Outcomes

##### MR-PREG Collaboration

The MR-PREG Collaboration currently includes data on up to 714,899 women for 35 pregnancy and perinatal outcomes^17^. MR-PREG includes GWAS summary data for these outcomes that we generated by pooling GWAS results from four core studies (the Avon Longitudinal Study of Parents and Children [ALSPAC], Born in Bradford [BiB], the Norwegian Mother, Father and Child Cohort Study [MoBa], and UKB), as well as GWAS data from a Finnish biobank (FinnGen) and published meta-analyses for specific outcomes. A description of these studies can be found in the Supplemental Information. In each GWAS, we have accounted for population structure by adjusting for principal components of ancestry and/or using mixed models. A more detailed description of the outcome definitions, exclusions, and genotypic data quality control conducted for each study and at the meta-analysis level can be found in the collaboration profile^17^. We use summary statistics for 19 out of the 35 pregnancy and perinatal outcomes, reflecting those we consider key safety outcomes. These are listed in **Table 1**.

**Table 1.** Adverse pregnancy and perinatal outcomes (APPOs) and underlying traits examined in this project.

| Type | Outcomes/Traits | Total sample size | Case number (%) |
| --- | --- | --- | --- |
| Pregnancy | Hypertensive disorders of pregnancy | 541,768 | 32,549 (6.0%) |
|  | Gestational hypertension | 527,932 | 20,777 (3.9%) |
|  | Pre-eclampsia | 671,992 | 19,408 (2.9%) |
|  | Gestational diabetes | 714,899 | 24,752 (3.5%) |
|  | Miscarriage | 486,217 | 89,086 (18.3%) |
|  | Recurrent miscarriage | 321,756 | 6,233 (1.9%) |
|  | Stillbirth | 207,670 | 6,331 (3.0%) |
| Delivery | Induction of labour | 95,239 | 14,495 (15.2%) |
|  | Premature rupture of membranes | 310,621 | 21,839 (7.0%) |
|  | Caesarean section | 233,969 | 33,909 (14.5%) |
|  | Spontaneous pre-term birth | 272,940 | 17,176 (6.3%) |
|  | Small for gestational age | 96,305 | 7,448 (7.7%) |
|  | Large for gestational age | 96,305 | 10,468 (10.9%) |
|  | Low Apgar score at 1 minute | 86,668 | 4,950 (5.7%) |
|  | Low Apgar score at 5 minutes | 74,368 | 876 (1.1%) |
|  | Postpartum haemorrhage* | 331,792 | 21,521 (6.5%) |
| Postnatal | NICU admission | 77,285 | 6,996 (9.1%) |
|  | Any Congenital anomalies | 74,701 | 3,569 (4.8%) |
| Continuous traits | Birth weight | 289,846 | N/A |
|  | Gestational age | 211,416 | N/A |

\* Postpartum haemorrhage GWAS is a publicly available GWAS and is not part of the MR-PREG meta-analyses.

##### Postpartum haemorrhage (PPH) GWAS meta-analysis by Westergaard et al., 2024

The PPH GWAS (N=302,894) conducted by Westergaard and colleagues^18^ included six cohorts of Western European ancestry: the Copenhagen Hospital Biobank Study on Reproduction (Denmark), Estonian Biobank (Estonia), FinnGen (Finland), deCODE genetics (Iceland), UKB (United Kingdom) and MoBa (Norway). GWAS from each cohort were combined using fixed-effects inverse variance weighted meta-analyses. The sample size for this dataset is also listed in **Table 1**.

#### Additional sources

##### UK Biobank Pharma Proteomics Project (UKB-PPP)

The UKB-PPP is a subset of the UKB cohort (N = 52,365) that was created to characterise the plasma proteomic profile. The protein apo(a) (encoded by *LPA*), along with 2,940 other blood plasma protein analytes, were measured using the antibody-based proximity extension assay by Olink (https://olink.com). The sample selection procedure and quality control by the UKB and the PPP consortium were previously described^19^. GWAS effect estimates for this dataset in the Olink NPX (Normalized Protein eXpression) values are a proprietary unit of relative (as opposed to absolute) protein abundance, expressed on a log2 scale. The effect size of MR estimates stemming from this dataset therefore have limited interpretability on their own and are meant to test for consistency in the directionality of estimates across exposure datasets.

##### GWAS meta-analysis by Mack et al., 2017

Mack and colleagues conducted a GWAS meta-analysis (N = 13,781) of Lp(a) levels in five different European cohorts: the CoLaus study (Switzerland), the National Heart, Lung, and Blood Institute (NHLBI) Family Heart Study (USA), Cooperative Health Research in the Region of Augsburg (in German: Kooperative Gesundheitsforschung in der Region Augsburg) KORA F3 and F4 (Germany), Cardiovascular Risk in Young Finns Study (Finland), and (Salzburg Atherosclerosis Prevention Program in Subjects at High Individual Risk (SAPHIR) (Austria). All Lp(a) measurements and determination of apo(a) isoforms were conducted in the same laboratory for all participating studies. Lp(a) was measured in mg/dl and adjusted for apo(a) isoform, using ELISA and immunoblotting, respectively. The MR estimates using this dataset were scaled to trait difference or odds ratio per 97.65 mg/dL decrease in Lp(a) to make them comparable to the 210 nmol/L scale. A more detailed description of the cohorts and the methods used can be found in the manuscript^20^.

### Main analysis

All analyses were undertaken using R (version 4.3.3) and Python 3 (version 3.5.2) and *P* values were adjusted for multiple comparisons using 5% false discovery rate correction (FDR) using the Benjamini & Hochberg (1995) method. Both raw and FDR-corrected *P* values are presented in the analysis outputs (in the **Supplemental Tables**) for transparency and to aid interpretation. Although assumed by FDR correction, most of the outcomes considered in this study are not independent from one another, making this a conservative approach.

#### MR assumptions

As with other statistical approaches, the validity of MR results relies on certain assumptions, the core ones can be summarized as follows:

I. Relevance: the instrument is statistically strongly associated with the exposure of interest in the relevant population (pregnant women, in this study).
II. Independence: there should be no confounding between the instrument and outcome.
III. Exclusion restriction criteria: any effect of the instrument on the outcome is fully mediated by the exposure.

The relevance assumption can be formally tested, whilst violations of the independence and exclusion restriction assumptions can be explored in sensitivity analyses but not conclusively refuted^21^.

#### Generalized inverse variance weighted (gIVW) MR

The gIVW MR method is an extension of the IVW method which allows for the inclusion of moderately correlated genetic instruments by accounting for their correlation with each other. In short, weighted generalized linear regression was performed using a weighting matrix based on the standard errors of the IV associations with the outcome and the pairwise correlations between variants^22,23^. The correlations between variants were estimated based on a reference panel of 10,000 randomly selected unrelated White European individuals from the UK Biobank.

#### Transformation of MR estimates

The direction of MR estimates was reversed to reflect pharmacological modulation (i.e. LPA perturbation to mimic lowering of Lp(a)). Results are presented on two scales: odds ratios (binary traits) or mean differences (continuous traits) per 1 nmol/L and per 210 nmol/L reduction in Lp(a). Phase 2 Pelacarsen studies reported a median baseline Lp(a) of 235 nmol/L and average reductions of ∼80%, meaning a 90% reduction (≈210 nmol/L) is a plausible and clinically meaningful response. Other therapeutics in development, including Olpasiran^7^, Zerlasiran^8^, and Lepodisiran^6^, have shown mean Lp(a) reductions of 90–95%, making this scale relevant for this drug class.

### Sensitivity analyses

#### Instrumenting Lp(a) with alternative datasets

To increase confidence that the findings in the primary analysis are not driven by biases specific to the measurement method used in the UKB, we repeated the main analysis with Lp(a) or apolipoprotein(a) summary data from two alternative GWAS studies, UKB-PPP and Mack et al, 2017, each using a different detection method as described in the Data Sources section.

#### Adjusting for fetal genetic effects

This study focuses on maternal genetic effects. However, as the risk of adverse pregnancy outcomes can be influenced by fetal genotype, it is important to account for it in MR analyses. We achieved this by repeating the MR with SNP-outcome associations that have been adjusted for fetal genotype (also referred to as conditional maternal effects) in a weighted linear model, as implemented by the DONUTS ^24^ package in R (v1.0.0). Likewise, we also investigated total and direct fetal genetic effects (adjusted for maternal genotype) to aid our interpretation of our findings. The importance and rationale of these analyses are described in more detail in the MR-PREG collaboration profile^17^.

#### Identifying outlying studies with leave-one-out of studies

To check whether any one study has a predominant effect, we performed MR on each single study contributing with outcome data and then meta-analysed all studies except one, repeating this to exclude each study in turn.

#### Genetic colocalisation at target locus for LD confounding

We performed genetic colocalisation of the association of genetic variants with Lp(a) levels and with each APPO at the *LPA* locus to explore whether MR results are driven by a shared causal variant for these two traits, or by confounding due to linkage disequilibrium. Genetic colocalisation consists of a Bayesian test of whether two traits, which may have an independent GWAS signal in the same locus, share a causal variant. If so, the likelihood of these two traits being causally linked by a biological pathway is greater. Colocalisation thus estimates the posterior probability of the following hypotheses (biomarker and disease example):

- PP.H0: neither trait has a genetic association in the locus.
- PP.H1: only biomarker trait has a genetic association in the locus.
- PP.H2: only disease trait has a genetic association in the locus.
- PP.H3: both traits have a genetic association in the locus, but with different causal variants.
- PP.H4: both traits have a genetic association in the locus, and share a single causal variant.

The estimation of posterior probabilities is based on the input data (genetic association estimates), as well prior probabilities: (i) p1: the variant is associated with biomarker levels, (ii) p2: the variant is associated with disease risk, and (iii) p12: the variant is associated with both traits. These were set at the defaults of 1×10^-4^, 1×10^-4^ and 1×10^-5^, respectively.

Colocalisation between Lp(a) and APPOs were tested using the coloc method^25^ and package (version 5.2.3) in R within the same window as the window of IV selection i.e. 500kb flanking the gene.

#### Accounting for sample overlap

To increase confidence that results are not influenced by sample overlap we employed the MRlap method which corrects for biases stemming from sample overlap, weak instruments and winner’s curse^26^. Because this analysis requires full GWAS summary statistics, we used the publicly available Lp(a) GWAS from the UKB by Sinnott-Armstrong and colleagues (N = 253,570) ^27^. The differences between this GWAS and the one by Shi et al., 2024, used for the IVs, are (i) the latter used an extended Lp(a) measurements dataset which included measurements outside of the assay’s reportable range and (ii) the latter included individuals self-reporting as European/Caucasian whilst Sinnott-Armstrong and colleagues included only those calculated to be White British.

#### Positive control MR

To investigate the credibility of the selected SNPs as instruments to estimate the effect of Lp(a), we repeated the main MR analysis using coronary artery disease as a positive control outcome, given the well-established causal role of Lp(a) on coronary artery disease risk^28^. The SNPs were extracted from the CARDIoGRAMplusC4D coronary artery disease GWAS (Nikpay *et al*., 2015, Open GWAS ID ieu-a-7, last accessed 2025DEC17) on OpenGWAS^30,31^, using SNPs at LD > 0.8 as proxies if needed.

#### Exploring Lp(a) at genetically-predicted very low or null levels

These analyses were performed using individual-level data from a White European UKB sample (itself part of MR-PREG), comprising up to 198,854 women (after latest withdrawals of consent as of October 2025). The analyses were performed on the UK Biobank Research Analysis Platform hosted by DNAnexus. These are described in detail in the Supplemental Information.

## Results

### Main analysis

Our main MR findings did not support a causal effect of Lp(a) lowering on APPOs (**Figure 2**). First, across all outcomes, MR estimated null to modest effects on outcomes per 210 nmol/L genetically predicted Lp(a) decrease. Second, for most outcomes, except gestational age, stillbirth and congenital malformations, 95% confidence intervals included the null value. Of these, the strongest trend is for genetically predicted lower Lp(a) to be associated with lower risk of congenital malformations. Third, none of the estimates passed multiple testing burden correction.

**Figure 2:**
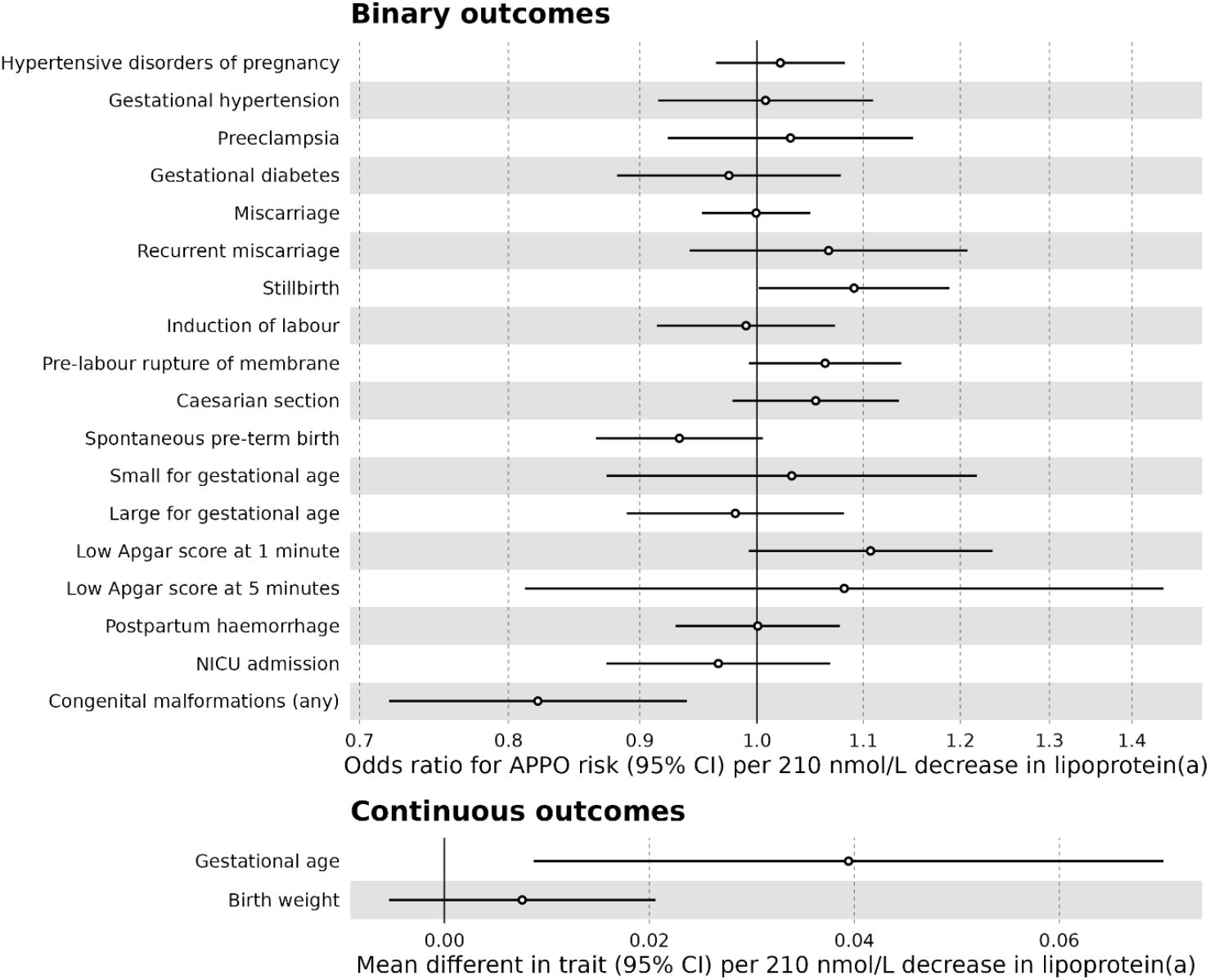
Mendelian randomization estimates for the estimated effect of Lp(a) lowering on adverse pregnancy and perinatal outcomes for a genetically-predicted decrease of Lp(a) by 210 nmol/L. Results are expressed as odds ratio (binary outcomes) or mean difference in trait (continuous outcomes) per 210 nmol/L decrease in Lp(a) resulting from genetically-instrumented downregulation of the target (LPA). Point estimates and 95% confidence intervals are represented by circles and bars, respectively. Open circles indicate FDR-corrected P>0.05. No examined trait had FDR-corrected P < 0.05. Gestational age is in weeks and birth weight is in Z units.

For several outcomes, MR estimates were close to null, with confidence intervals spanning both directions of effect: gestational hypertension, gestational diabetes, preeclampsia, NICU admission, miscarriage, small for gestational age, large for gestational age, induction of labour, postpartum haemorrhage, and caesarean section. For other outcomes, MR estimated small positive effects for a genetically-predicted decrease by 210 nmol/L with confidence intervals, often including the null: gestational age (β = 0.039 weeks per SD, 95% CI: 0.016, 0.056), recurrent miscarriage (OR = 1.066, 95% CI: 0.941, 1.208), pre-labour rupture of membranes (OR = 1.063, 95% CI: 0.993, 1.138), hypertensive disorders of pregnancy (OR = 1.049, 95% CI: 0.985, 1.117), stillbirth (OR = 1.091, 95% CI: 1.002, 1.188), low Apgar score at 1 minute (OR = 1.107, 95% CI: 0.993, 1.235), and low Apgar at 5 minutes (OR = 1.081, 95% CI: 0.812, 1.440). Finally, small negative associations (lower Lp(a) associated with lower odds of the adverse outcome) were observed for congenital malformations (OR = 0.821, 95% CI: 0.719, 0.939) and pre-term birth (OR = 0.933, 95% CI: 0.865, 1.005).

The size of the estimates when considering a 1 nmol/L decrease in Lp(a) levels is shown in **Figure S1**. The full output of the main analysis is shown in **Table S3**.

### Sensitivity analyses

#### Instrumenting Lp(a) with alternative datasets

When considering alternative exposure GWAS datasets, each featuring a different assaying method, the directionality of the MR estimates was consistent across the three analyses (**Figure 3, Table S4**). The effect sizes of Olink UKB-PPP-based MR estimates are not expressed in absolute abundance units and thus not comparable in magnitude to the other two datasets, which are in the nmol/L (main) and mg/dL units and scaled to reflect roughly the effects of equivalent changes in Lp(a) levels. When comparing the MR estimates using the Mack et al., 2017 GWAS with the main results, the predicted effects of Lp(a) lowering were larger and did not span the null for several outcomes: caesarian section, NICU admission, stillbirth, spontaneous preterm birth, miscarriage, congenital malformations, birth weight and gestational age. However, in all instances, the 95% confidence interval of the Mack et al., 2017 MR estimates spanned the 95% confidence interval of the main MR estimates, indicating little evidence of a difference between effect sizes. When comparing with the Olink UKB-PPP GWAS MR estimates, the effects of lower Lp(a) were generally directionally consistent with the ones from the main analysis, with the estimates for preterm birth, preeclampsia and induction of labour not spanning the null.

**Figure 3:**
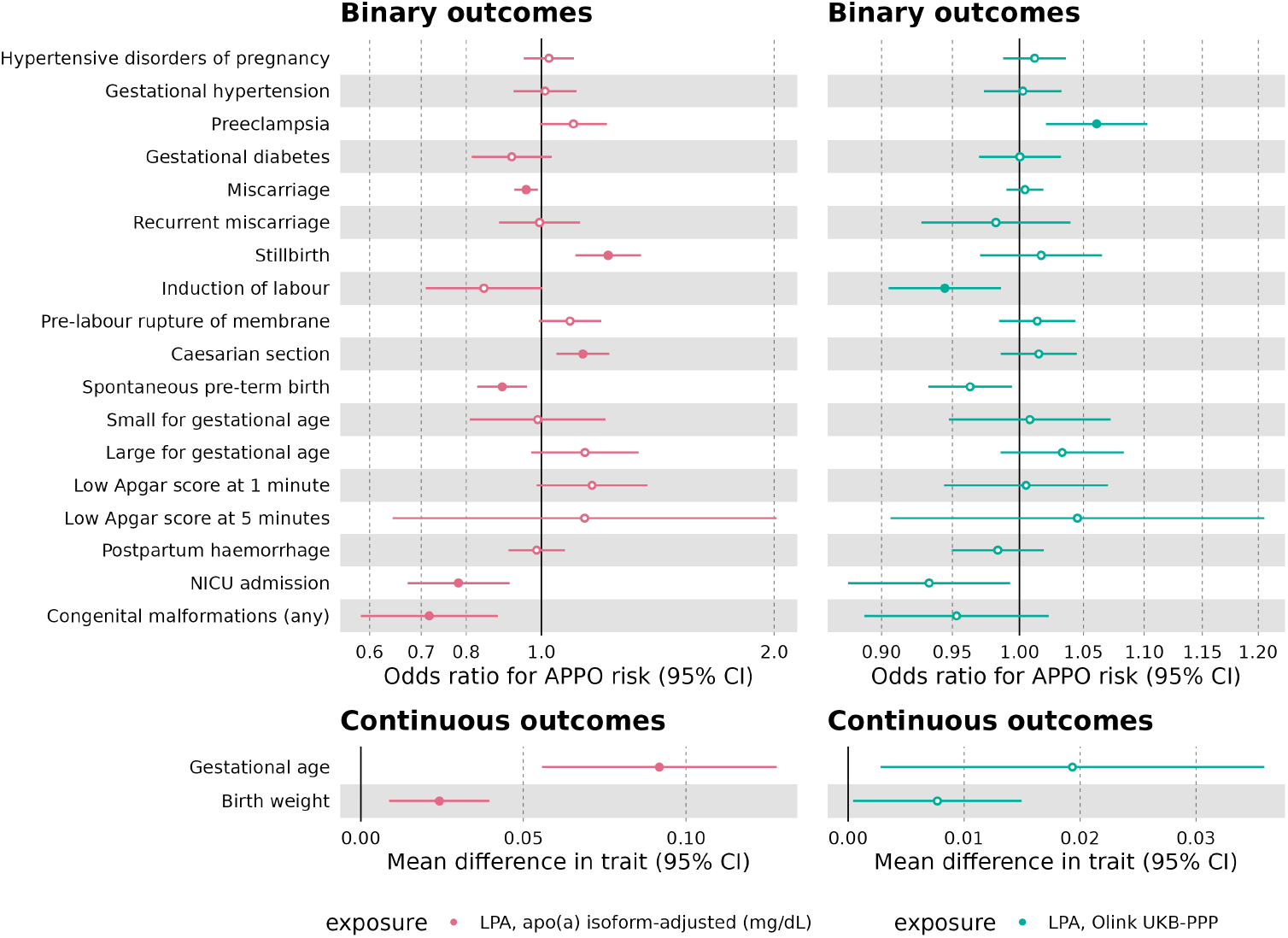
Mendelian randomization estimates for the estimated effect of Lp(a) lowering on adverse pregnancy and perinatal outcomes using alternative exposure datasets. Results are expressed as odds ratio (binary outcomes) or mean difference in trait (continuous outcomes) for every 97.65 mg/dL (which is roughly equivalent to 210nmol/L) (Mack et al., 2017 GWAS, left hand side) decrease in Lp(a) resulting from genetically-instrumented downregulation of the drug target gene (LPA). Results using the Olink (right hand side) UKB-PPP dataset are in NPX units which are in a log2 scale; these reflect relative as opposed to absolute abundance. Point estimates and 95% confidence intervals are represented by circles and bars, respectively. Open circles indicate FDR-corrected P > 0.05. Gestational age is in weeks and birth weight is in Z units.

#### Adjusting for fetal genetic effects

MR analyses were repeated using marginal and conditional (fetal-adjusted) outcome GWAS estimates from our weighted linear models. We observed no major differences in direction or effect size between unadjusted and adjusted MR estimates besides some expected widening of 95% confidence intervals for fetal-adjusted estimates, such as with gestational diabetes (**Figure 4, Table S5**). It is worth noting the unadjusted MR estimates were very similar but not identical to main MR estimates as the marginal outcome GWAS association estimates are an output from the WLM analysis and the number of SNPs included may differ slightly. Additionally, we could not perform these analyses for pregnancy loss outcomes (miscarriage, recurrent miscarriage and stillbirth) due to the absence of fetal genotype data for those pregnancies.

**Figure 4:**
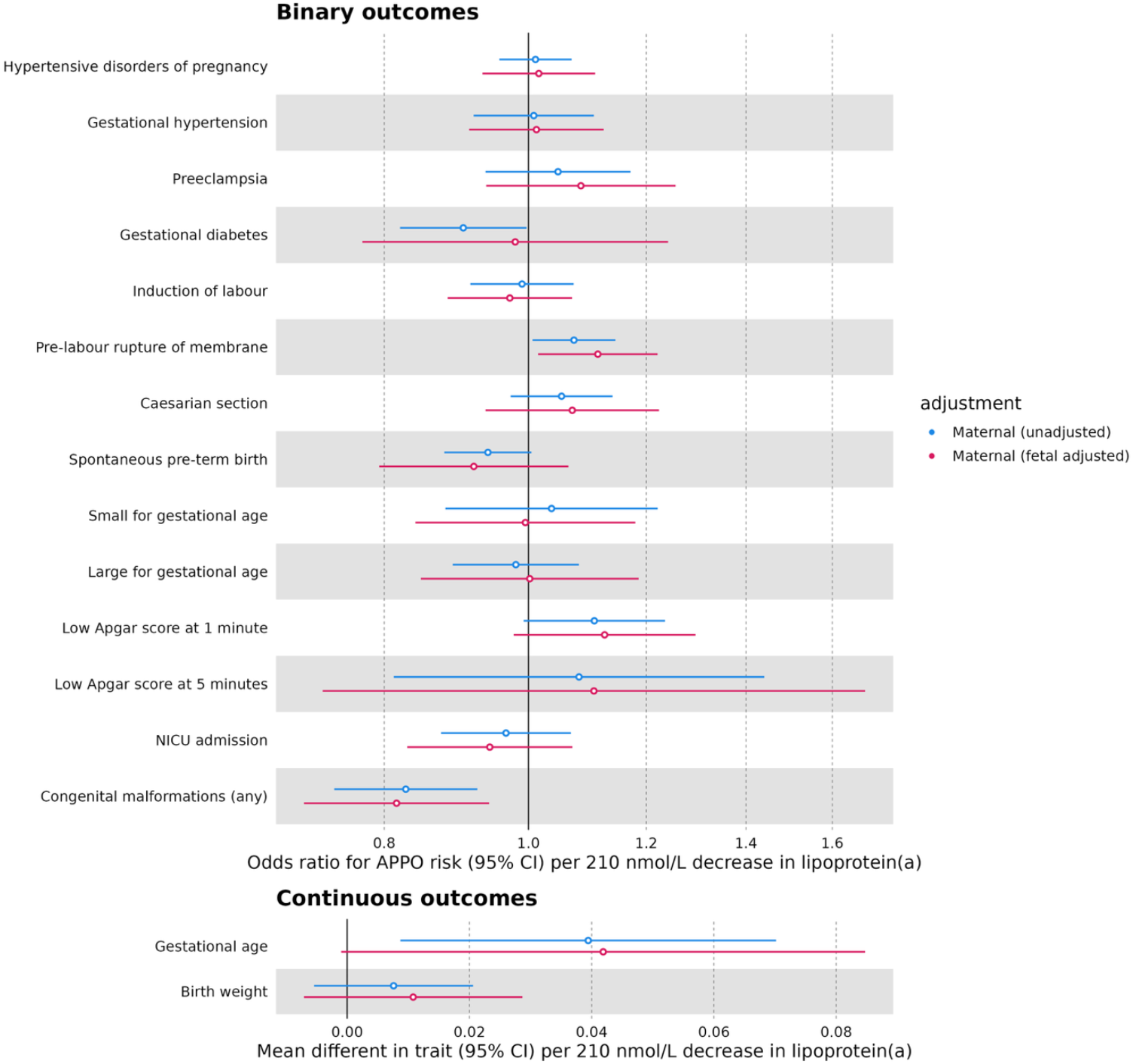
Mendelian randomization estimates for the estimated effect of Lp(a) lowering on adverse pregnancy and perinatal outcomes, unadjusted and adjusted for fetal genotype. Results are expressed as odds ratio (binary outcomes) or mean difference in trait (continuous outcomes) for every 210 nmol/L decrease in Lp(a) resulting from genetically-instrumented downregulation of the drug target gene (LPA). Point estimates and 95% confidence intervals are represented by circles and bars, respectively. Open circles indicate FDR-corrected P < 0.05. Gestational age is in weeks and birth weight is in Z units.

#### Identifying outlying studies with leave-one-out of studies

When performing leave-one-out of studies contributing with outcome data, the estimated effects of Lp(a) lowering were mostly consistent in magnitude and direction of effect (**Figure 5, Table S6**). In a small number of instances where removing a study reversed the direction of effect, the 95% confidence intervals of the MR estimate were widened and/or spanned 95% confidence interval of the meta-analysis MR estimate, and were the result of removing the largest contributing study (e.g. Low Apgar score at 5 minutes after removing MoBa and stillbirth after removing UKB). This analysis could not be performed for congenital anomalies as this outcome was only made up by the MoBa cohort, nor for PPH as we did not have access to the study-specific estimates for the GWAS meta-analysis ^18^.

**Figure 5:**
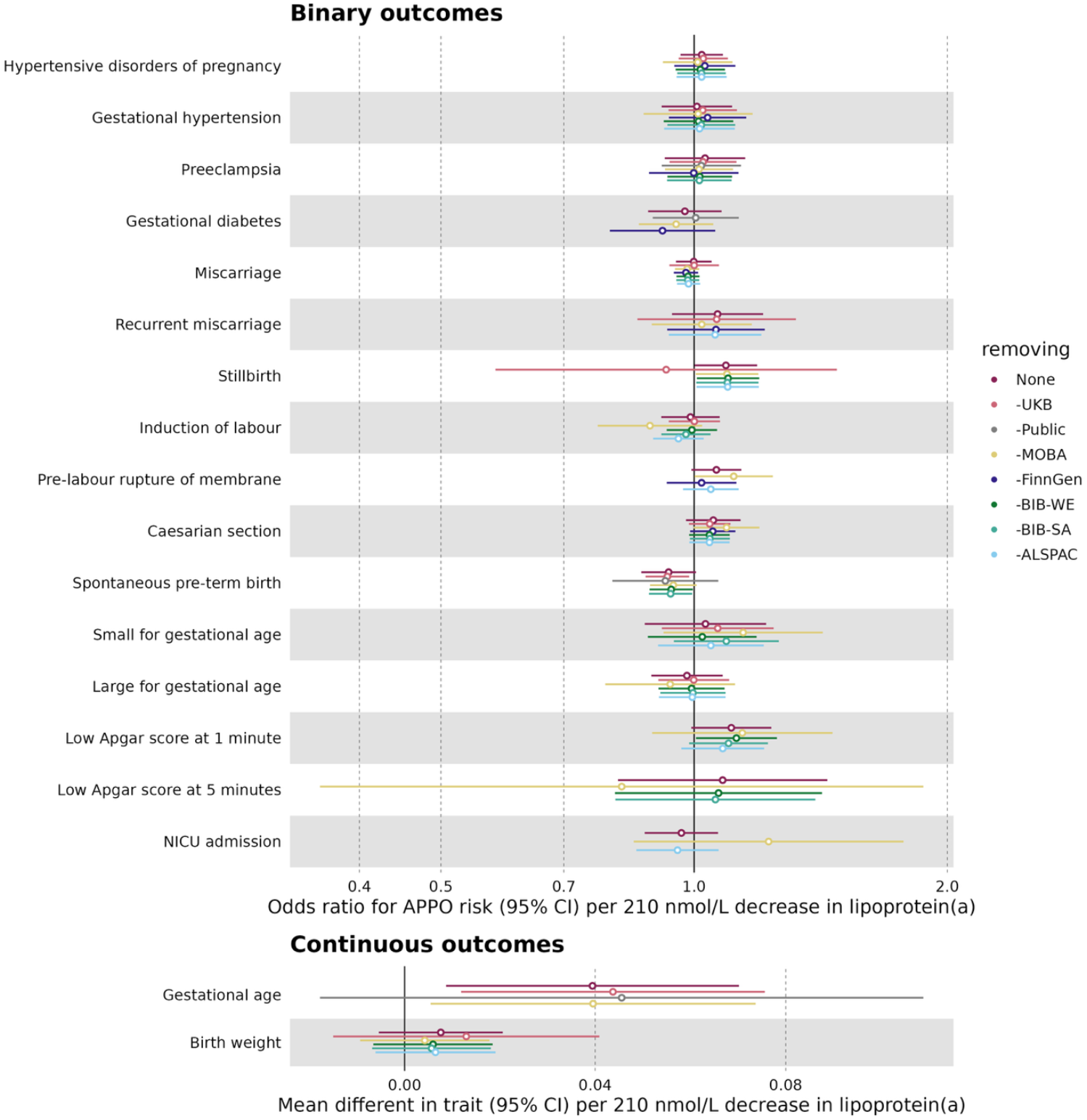
Leave-one-study-out Mendelian randomization estimates for the estimated effect of Lp(a) lowering on adverse pregnancy and perinatal outcomes. Results are expressed as odds ratio (binary outcomes) or mean difference in trait (continuous outcomes) for every 1 nmol/L decrease in Lp(a) resulting from genetically-instrumented downregulation of the drug target gene (LPA), after removing the contribution of the study specified in the label. Point estimates and 95% confidence intervals are represented by circles and bars, respectively. Open circles indicate FDR-corrected P > 0.05. Gestational age is in weeks and birth weight is in Z units.

#### Genetic colocalization

The genetic colocalization analysis showed no evidence of a shared causal variant between Lp(a) levels and APPOs at the *LPA* locus (PP.H_4_ = 0.01–0.11, **Table S7**). The posterior probabilities were indicative of no detection of causal variants for the APPOs, reflected by high probabilities for variants affecting only Lp(a) levels within the *LPA* locus (PP.H_1_ = 75–94%) as opposed to high probabilities for the traits having distinct causal variants in the locus (PP.H_3_ = 4–24%). An exception was birthweight, which showed modest evidence for distinct variants (PP.H_1_ = 52%; PP.H_3_ = 48%). These findings may indicate limited power to detect causal variants for the outcomes or a true absence of shared causal variants.

#### Accounting for sample overlap

We performed MRlap estimating the effect of causal effect Lp(a) on each of the APPOs, adjusting for any potential sample overlap between the exposure and outcome datasets. This analysis showed no evidence of a difference in effect estimates between the observed and the corrected estimates (**Table S8**), suggesting the partial sample overlap between the exposure and (some) outcome GWAS were not a driver of our results.

#### Positive control MR

In a positive control analysis, to assess the relevance of the selected instruments, we found robust evidence of a causal effect between genetically-predicted Lp(a) lowering and a lower risk of coronary artery disease (**Table S9**, OR = 0.55 per 210 nmol/L decrease in Lp(a), 95% CI: 0.46, 0.65).

#### Exploring Lp(a) at genetically-predicted very low or null levels

Across Lp(a) genetic score quintiles, associations with adverse pregnancy and perinatal outcomes were generally imprecise and showed no consistent trends (**Figure S4, Table S13**). Similarly, analyses of individuals homozygous for LoF *LPA* variants (akin to *LPA* knockouts) showed no robust associations with any outcomes, though estimates were limited by small numbers and wide confidence intervals (**Figure S5, Table S17**). A positive-control analysis of coronary artery disease suggested a protective direction of effect for loss of function of *LPA* but was too imprecise to confirm.

## Discussion

Across 20 outcomes, we found no robust evidence that a genetically predicted reduction in Lp(a) levels adversely affects pregnancy or perinatal health. Most estimates were modest in size with 95% confidence intervals excluding large effects despite being scaled to a large reduction of Lp(a) by 210 nmol/L. For comparison, we used CAD as a positive control and confirmed a substantially stronger effect on CAD risk than in any of the APPOs, with precise confidence intervals excluding the null.

Multiple sensitivity analyses did not indicate substantial bias. Results were consistent across alternative Lp(a) datasets, an apo(a) isoform-adjusted ELISA assay or a proteomics assay by Olink, indicating our main MR findings are not influenced by Lp(a) measurement methodology. We also adjusted for potential fetal genetic effects, which would violate the exclusion restriction assumption when investigating the effect of modulating maternal, as opposed to fetal, Lp(a) levels. Our results indicate that our main MR estimates were not influenced by fetal genetics; they also suggest a lower likelihood of the risk of these outcomes changing as a result of a Lp(a)-modulating drug crossing the placenta. Similarly, our leave-one-out analyses indicated that none of the contributing cohorts within MR-PREG were disproportionately responsible for our results.

We additionally probed whether null or very low Lp(a) levels, which could be pharmacologically induced by very effective Lp(a) lowering pharmacodynamics, were associated with increased risk of adverse outcomes using LoF variants in the *LPA* gene as a natural human knockout model. We found no evidence that *LPA* knockouts have a higher risk of the available APPOs. This provides a complementary line of evidence to the main MR analyses, which assume linearity and, therefore, may miss effects driven by extreme values. However, these results should be interpreted as exploratory, as power was limited; even for the positive-control outcome, CAD, the estimate showed the expected protective direction of effect^32^ but with a confidence interval that marginally included the null.

Although recent work has improved understanding of Lp(a) biology, major gaps remain^33^, and its role in pregnancy and development is even less well understood. Most studies over the last four decades report that Lp(a) increases during pregnancy^34–38^, yet it is still unclear whether these trajectories are altered in pregnancies complicated by adverse outcomes as previous studies have relied on observational comparisons that cannot distinguish causal effects from secondary physiological changes.

Among APPOs, preeclampsia has been most investigated to date as endothelial dysfunction and thrombosis are hallmarks with which Lp(a) has been linked in atherosclerotic disease; however, findings have been mixed^36,39–43^. A previous MR study reported no evidence of a causal effect of Lp(a) levels on preeclampsia risk^44^. Building on this limited evidence base, our results also did not support a causal role in preeclampsia. Notably, even in PPH, where the potential added risk from impaired clot formation contrasts with the prothrombotic nature of preeclampsia, evidence was not supportive of a causal role by Lp(a). Evidence for other outcomes is sparse, with some small retrospective studies reporting increased levels in those with history of unexplained recurrent miscarriage or stillbirth^45,46^. Our MR estimates corresponding to a large reduction in Lp(a) did not support an effect on miscarriage. The results were less conclusive for the rarer outcomes, for which we observed a slightly elevated risk with wide confidence intervals (recurrent miscarriage) or that did not withstand correction for multiple testing (stillbirth). This might have been a result of lack of power, so we cannot rule out the existence of modest but still clinically meaningful effects.

Overall, these findings can provide some early indication that substantial lowering of Lp(a) levels does not associate strongly with pregnancy and perinatal risks, especially for more common outcomes such as hypertensive disorders of pregnancy, gestational diabetes, and miscarriage. Upon the introduction of novel nucleic acid therapies targeting Lp(a) into preventative cardiovascular care, these findings may guide early risk-benefit discussions for individuals of childbearing potential. Nonetheless, these findings in isolation are insufficient to inform indications or guide decisions on whether to commence or halt therapy around the period of conception. Further studies will be crucial to better inform on the reproductive and developmental safety of these therapies. Additional strategies may evaluate apo(a) isoforms and variants located in the KIV-2 domains, diverse ancestral backgrounds, the genetic basis of Lp(a) levels throughout the gestational period, and evidence from databases linking prescription data to EHR and maternity records.

### Strengths and limitations

Human genetics provides a powerful framework for identifying mechanism-based adverse effects prior to clinical testing^47^. In this study, we benefited from the particularly strong biological alignment between the genetic instrument and the therapeutic mechanism. Lp(a) levels are largely determined by genetic variation in the *LPA* locus, with our instruments explaining up to 64% of Lp(a) variance. The *LPA* gene is almost exclusively expressed in the liver^48^, the same organ targeted by both ASO and siRNA therapies acting on *LPA* mRNA^49^. This increases confidence in the relevance and validity of our instruments to mimic pharmacological perturbation and supports the plausibility of the MR assumption of gene–environment equivalence.

A key strength of this study is the depth and breadth of datasets and outcomes assessed. We examined a wide range of APPOs with large sample sizes, most having substantial case numbers. We also disentangled maternal and fetal effects, which are an important but often overlooked aspect of intergenerational MR research, and conducted a wide range of sensitivity analyses which generally supported the robustness of the main results. Finally, complementary analyses in human ‘knockouts’ provided an additional line of evidence at the extreme low end of the Lp(a) distribution.

Some limitations in this study relate to the data sources. Samples sizes and case numbers were limited for rarer outcomes, reducing statistical power and increasing uncertainty. Furthermore, the studies included in the MR-PREG mostly include individuals of White European ancestry, with only a small South Asian sample, limiting generalisability to other ancestry groups. Finally, outcomes were limited to the perinatal period, limiting assessment of long-term maternal or offspring outcomes. For congenital malformations, data were available only for cases identified at birth in MoBa, not capturing malformations leading to pregnancy loss or those detected later in life, or determining whether specific types drive any potential associations.

Other limitations relate to the independence and exclusion restriction MR assumptions. While we conducted extensive sensitivity analyses to assess their plausibility, they cannot be empirically verified. We cannot exclude the possibility that the select cis *LPA* variants influence the expression of other genes, a violation of the exclusion restriction assumption. Because fetal genotype data is not available for pregnancy loss phenotypes, bias from fetal effects could not be assessed for miscarriage and stillbirth.

Finally, there are caveats in translating drug-target MR findings to pharmacological interventions. We assumed that genetic regulation of Lp(a) levels during pregnancy is similar to that observed in a Lp(a) GWAS of middle-aged women and men due to lack of available data. MR typically reflects long-term changes in the exposure of interest and therefore cannot disentangle effects occurring within specific gestational windows. Moreover, our main MR analyses assume a linearity. We conducted individual-level analyses to explore non-linear effects; however, these were underpowered. The methodology used throughout this study can only be used to estimate biological effects from perturbing a target and cannot capture drug-specific pharmacokinetic or pharmacodynamic properties that arise from delivery mechanisms or chemical properties.

## Conclusions

In summary, we do not find robust evidence of a causal effect of Lp(a) lowering on the assessed APPOs. Multiple sensitivity analyses reinforce these findings, reducing concerns about potential biases such as fetal genetic effects, dominance of a single study in meta-analyses, sample overlap, or confounding by linkage disequilibrium. Taken together, these results offer initial reassurance as Lp(a)-lowering therapies emerge, particularly for individuals with inherited high Lp(a) who may be considered for preventive treatment at childbearing age. However, modest effects, particularly for rarer outcomes such as stillbirth, cannot be ruled out and will need to be addressed in future genetic studies with larger sample sizes. Finally, interpretation of these findings relies on the validity of MR assumptions, particularly those specific to when extrapolating to pharmacologic modulation during pregnancy. These considerations should guide any clinical translations of our results.

## Data Availability

The ALSPAC access policy that describes the proposal process in detail including any costs associated with conducting research at ALSPAC, which may be updated from time to time (https://www.bristol.ac.uk/media-library/sites/alspac/documents/researchers/data-access/ALSPAC_Access_Policy.pdf). Data is available upon request from Born in Bradford (https://borninbradford.nhs.uk/research/how-to-access-data/). Data from MoBa are available from the Norwegian Institute of Public Health after application to the MoBa Scientific Management Group (https://www.fhi.no/en/op/data-access-from-health-registries-health-studies-and-biobanks/data-access/applying-for-access-to-data/). Researchers can apply for access to the UK Biobank data via the Access Management System (AMS) (https://www.ukbiobank.ac.uk/enable-your-research/apply-for-access).
All code used to conduct analyses and generate figures can be found at https://github.com/helenaurquijo/lpa_in_pregnancy (release v0.1.1).

https://www.fhi.no/en/op/data-access-from-health-registries-health-studies-and-biobanks/data-access/applying-for-access-to-data/

https://www.ukbiobank.ac.uk/enable-your-research/apply-for-access

https://github.com/helenaurquijo/lpa_in_pregnancy

## Abbreviations

ALSPAC: Avon Longitudinal Study of Parents and Children
AMS: Access Management System
APPO: Adverse Pregnancy and Perinatal Outcome
ASCVD: Atherosclerotic Cardiovascular Disease
ASO: Antisense Oligonucleotide
BiB: Born in Bradford
CAD: Coronary Artery Disease
CARDIoGRAMplusC4D: Coronary Artery Disease Genome-wide Association consortium
CI: Confidence Interval
ELISA: Enzyme-Linked Immunosorbent Assay
FDR: False Discovery Rate
GWAS: Genome-Wide Association Study
IV: Instrumental Variable
KIV-2: Kringle
IV: type 2 repeat
LD: Linkage Disequilibrium
LDL: Low-Density Lipoprotein
LoF: Loss of Function
Lp(a): Lipoprotein(a)
LPA: Apolipoprotein(a) gene
MAF: Minor Allele Frequency
MoBa: Norwegian Mother, Father and Child Cohort Study
MR: Mendelian Randomization
MR-PREG: Mendelian Randomization in Pregnancy Collaboration
NICU: Neonatal Intensive Care Unit
NIPH: Norwegian Institute of Public Health
NPX: Normalized Protein eXpression
OR: Odds Ratio
PCSK9: Proprotein Convertase Subtilisin/Kexin Type 9
PIP: Posterior Inclusion Probability
PLG: Plasminogen gene
PPH: Postpartum Haemorrhage
QC: Quality Control
RCT: Randomized Controlled Trial
siRNA: Small Interfering RNA
UKB: UK Biobank
UKB-PPP: UK Biobank Pharma Proteomics Project

## Data and code availability

The ALSPAC access policy that describes the proposal process in detail including any costs associated with conducting research at ALSPAC, which may be updated from time to time (https://www.bristol.ac.uk/media-library/sites/alspac/documents/researchers/data-access/ALSPAC_Access_Policy.pdf). Data is available upon request from Born in Bradford (https://borninbradford.nhs.uk/research/how-to-access-data/). Data from MoBa are available from the Norwegian Institute of Public Health after application to the MoBa Scientific Management Group (https://www.fhi.no/en/op/data-access-from-health-registries-health-studies-and-biobanks/data-access/applying-for-access-to-data/). Researchers can apply for access to the UK Biobank data via the Access Management System (AMS) (https://www.ukbiobank.ac.uk/enable-your-research/apply-for-access).

All code used to conduct analyses and generate figures can be found at https://github.com/helenaurquijo/lpa_in_pregnancy (release v0.1.1).

## Acknowledgements

This work was carried out using the computational facilities of the Advanced Computing Research Centre, University of Bristol (http://www.bristol.ac.uk/acrc/).

ALSPAC: We are extremely grateful to all the families who took part in this study, the midwives for their help in recruiting them, and the whole ALSPAC team, which includes interviewers, computer and laboratory technicians, clerical workers, research scientists, volunteers, managers, receptionists and nurses.

BiB: BiB is only possible because of the enthusiasm and commitment of the Children and Parents in BiB. We are grateful to all the participants, teachers, school staff, health professionals and researchers who have made BiB happen.

MoBa: This research has been conducted using MoBa data using application number 2552. MoBa is supported by the Norwegian Ministry of Health and Care services and the Ministry of Education and Research. We are grateful to all the participating families in Norway who take part in this on-going cohort study. We thank the Norwegian Institute of Public Health (NIPH) for generating high-quality genomic data. This research is part of the HARVEST collaboration, supported by the Research Council of Norway (#229624). We also thank the NORMENT Centre for providing genotype data, funded by the Research Council of Norway (#223273), South East Norway Health Authority and KG Jebsen Stiftelsen. We further thank the Center for Diabetes Research, the University of Bergen for providing genotype data and performing QC and imputation of the data funded by the ERC AdG project SELECTionPREDISPOSED, Stiftelsen Kristian Gerhard Jebsen, Trond Mohn Foundation, the Research Council of Norway, the Novo Nordisk Foundation, the University of Bergen, and the Western Norway health Authorities (Helse Vest).

FinnGen: The authors thank the FinnGen investigators for sharing their summary-level data.

UK Biobank: We would like to thank all the participants of UK Biobank for their vital contribution to the resource. This research has been conducted using the UK Biobank Resource under Application Number 23938.

