## Supplemental Materials for "Using human genetic variation to estimate the effect of lipoprotein(a) lowering on pregnancy outcomes"

5- Translational Medicine, Biomedical Research, Novartis, Prague, Czechia.

6- Centre for Fertility and Health, Norwegian Institute of Public Health, Oslo, Norway.

7- NIHR Bristol Biomedical Research Centre, University of Bristol, Bristol, UK.

* These authors contributed equally.

**Corresponding Author:**

Helena Urquijo

MRC Integrative Epidemiology Unit

University of Bristol

Augustine's Courtyard

Orchard Lane

BS1 5DS, Bristol

United Kingdom

### Supplemental Information

#### Description of MR-PREG contributing cohorts

In this study, we used data from the MR-PREG collaboration (release 4), including up to 671,922 women from four cohort studies:  Avon Longitudinal Study of Parents and Children (ALSPAC)[1,2]; Born in Bradford (BiB)[3]; the Norwegian Mother, Father and Child Cohort Study (MoBa)[4], UK Biobank (UKB)[5], and public data from FinnGen[6] and previous genome-wide association study (GWAS) metanalyses for pre-eclampsia[7], pre-term birth[8], gestational age[8] and gestational diabetes[9].

##### *ALSPAC: The Avon Longitudinal Study of Parents and Children*

ALSPAC is a prospective birth cohort that started recruiting pregnant women resident in the former county of Avon (centred around the city of Bristol, Southwest England), with delivery dates between April 1991 and December 1992. A total of 14,541 women (ALSPAC-G0) were enrolled during pregnancy (14,676 fetuses) and gave birth to 14,062 live children (ALSPAC-G1). 7 years after the initial recruitment, a further 913 eligible children were enrolled. The total sample size for analyses using any data collected after the age of seven is therefore 15,447 pregnancies (14,833 unique mothers), resulting in 15,658 fetuses. Further details can be found in the published cohort profiles[1,2]. Please note that the study website contains details of all the data that is available through a fully searchable data dictionary and variable search tool" and reference the following webpage: http://www.bristol.ac.uk/alspac/researchers/our-data/. Ethical approval for the study was obtained from the ALSPAC Ethics and Law Committee and the Local Research Ethics Committees.

##### *BiB: Born in Bradford*

Born in Bradford (BiB) is a prospective birth cohort that recruited women with expected delivery between March 2007 and December 2010. Most women were recruited at their oral glucose tolerance test (OGTT) at approximately 26–28 weeks' gestation, which is offered to all women booked for delivery at Bradford Royal Infirmary, except those with known diabetes. In BiB, most of the obstetric population consists of women of White British or Pakistani origin (together accounting for 81%, with the remaining women being of other ancestries).  A total of 12,453 women (13,776 pregnancies) were enrolled during pregnancy who gave birth to 13,858 live children. Full details of the study methodology were reported previously[3]. Ethical approval for the study was granted by the Bradford National Health Service Research Ethics Committee (ref 06/Q1202/48).

##### *MoBa: The Norwegian Mother, Father and Child Cohort Study*

The Norwegian Mother, Father and Child Cohort Study (MoBa) is a prospective birth cohort that recruited pregnant women from all over Norway from 1999-2008. From all pregnant women, 41% consented to participate. The cohort includes approximately 114,500 children, 95,200 mothers and 75,200 fathers. The establishment of MoBa and initial data collection was based on a license from the Norwegian Data Protection Agency and approval from The Regional Committees for Medical and Health Research Ethics. The MoBa cohort is currently regulated by the Norwegian Health Registry Act. Ethical approval for our study was obtained from The Regional Committees for Medical and Health Research Ethics (ref 2018/1256).

##### *UKB: UK Biobank*

All people in the UK National Health Service (NHS) registry aged between 40-69 years and living within approximately 25-mile radius from one of the 22 study centers were invited to participate in UK Biobank (UKB) between 2006-2010 (6, 12). In total, 503,325 adults (5.5% of the ~9.2 million invited) were recruited into the study. Ethical approval for UKB was obtained from the Northwest Multi-Centre Research Ethics Committee (MREC), and our study was performed under UKB application number 23938.

##### *FinnGen*

FinnGen is the national wide network of Finnish biobanks. At the time of writing, FinnGen included data from 453,733 individuals (254,618 females and 199,115 males) [12th data release (R12)]. The Coordinating Ethics Committee of the Helsinki and Uusimaa Hospital District has approved the FinnGen consortium (Nr HUS/990/2017), and the ethical approval of each individual study has been described in detail elsewhere (PMID: 32066667). The metadata from FinnGen used by the MR-PREG collaboration is publicly available at https://www.finngen.fi/en/access_results.

##### *Published GWAS metanalyses*

In this study, we used public genetic association data from the International Pregnancy Genetics (InterPregGen), the Early Growth Genetics (EGG) consortium and the GENetics of Diabetes in Pregnancy Consortium (GenDIP) consortium. InterPregGen is a GWAS meta-analysis for HDP, including 9,515 cases of preeclampsia and 157,719 controls (76% European and 24% Central Asian cases). The EGG consortium includes gestational duration-related traits includes GWAS of preterm birth (N = 18,797 cases and 260,246 controls), postterm birth (N = 15,972 cases and 115,307 controls) and gestational duration (N = 195,555) in European ancestry women. GenDIP has a GWAS meta-analysis of 5485 gestational diabetes cases and 347 856 controls[9].

#### Outcome definitions

All outcomes only include singleton pregnancies.

**Postpartum haemorrhage**[10]: Haemorrhage (>500 ml) following birth according to ICD10 code O72. Those with known coagulation disorders and having undergone a caesarean section were excluded.

An additional description of the study-specific data sources to define outcomes and of deviations from these outcome definitions are presented the MR-PREG cohort profile[11].

**Gestational age:** Gestational age in weeks, excluding non-live births, elective caesarean section and induced births.

**Spontaneous pre-term birth:** Birth at gestational age <=37 weeks. Controls were those with birth at gestational ages between 37 and 42 weeks. Non-live births, elective caesarean section and induced births were excluded.

**Preeclampsia:** Hypertensive disorder of pregnancy (as below) PLUS at least 1+ proteinuria occurring at the same time as the episodes of raised blood pressure. Controls were those with no HDP. Non-live births were excluded.

**Gestational hypertension**: Hypertension defined as systolic blood pressure >139mmHg OR diastolic blood pressure >89mmHg on at least 2 occasions after 20 weeks of gestation in women who had not previously been diagnosed with hypertension outside of pregnancy. The 'healthy' (coded 0) group includes those who had a previous diagnosis of hypertension prior to pregnancy. Non-live births were excluded.

**Hypertensive disorders of pregnancy (HDP):** Gestational hypertension OR preeclampsia. Non-live births were excluded.

**Miscarriage:** Loss of pregnancy until 20 weeks' gestation.

**Recurrent miscarriage** ≥ 3 miscarriages in the index and previous pregnancies. Individuals were excluded if they had history of menarche before 9 years or after 17 years of age, or of specific medical conditions.

**Stillbirth:** Loss of pregnancy between after 20 weeks' gestation.

**Gestational diabetes:** Diabetes mellitus in pregnancy, self-reported or from medical records [ICD-10 code O24]. Non-live births were excluded.

**Congenital anomalies (any):** Congenital malformations according to the Medical Birth Registry of Norway (MBRN). Non-live births were excluded.

**Induction of labour:** Labour was medically or surgically induced. Controls were those with no induced labour. Non-live births were excluded.

**Birth weight**: Weight at birth in SD units with z-scores transformed separately in males and females. Extracted from medical records or according to self-report in questionnaires. Non-live births and birth weight > 5 SD were excluded.

**Small for gestational age**: Birth weight-for-gestational age < 10th percentile. Controls were those on and above 10th percentile. Non-live births were excluded.

**Large for gestational age**: Birth weight-for-gestational age > 90th percentile. Controls were those on and below 90th percentile. Non-live births were excluded.

**Low Apgar score at 1 minute**: Birth with Apgar score < 7 at 1 minute. Controls were those with Apgar score >= 7. Non-live births were excluded.

**Low Apgar score at 5 minutes**: Birth with Apgar score < 7 at 5 minutes. Controls were those with Apgar score >= 7. Non-live births were excluded.

**NICU admission**: Births with admission to neonatal intensive care unit (NICU). Controls were those not admitted to NICU. Non-live births were excluded.

**Caesarian section:** Births occurring through caesarian section. Controls were those not born through caesarian section. Non-live births were excluded.

**Premature rupture of membranes:** Premature rupture of membranes occurring at any gestational age. Non-live births were excluded.

#### Exploring Lp(a) at genetically-predicted very low or null levels

To address added interest in thrombotic events due to the homology between apo(a) and plasminogen, these analyses also included venous and arterial thrombotic events during pregnancy as two additional outcomes. Venous thrombotic events were selected using the ICD10 codes [and rough ICD9 equivalents]: O22.2 [671.2], O22.3 [671.3], O22.5 [671.5], O22.9 [671.5, 671.9], O87.0 [671.2], O87.1 [671.4], O87.3 [671.5], O87.9 [671.5] and O88.2 [673.2]. These cover venous thromboembolic complications during pregnancy and the puerperium, including both superficial and deep vein thromboses, cerebral venous thrombosis, and unspecified venous issues. They also include obstetric embolisms, particularly blood-clot (pulmonary) embolisms. Arterial thrombotic events such as MI and stroke [674.0] were selected using O99.4 in ICD10 [648.6], which captures all circulatory conditions (I00-I99) during pregnancy that do not have a specific code.

All regression models included the first 40 genetic principal components (data field p22009) and genotyping batch (data field p22000) as covariates.

#### Lp(a) genetic score

We created a genetic score of Lp(a) using plink 1.9[12] as implemented in the ukbrapR R package (https://github.com/lcpilling/ukbrapR). The Lp(a) genetic score included only the cis variants in the *LPA* region and was weighted according to conditional estimates of the variants' effect on Lp(a) to account for correlation between variants. Independent SNPs were identified using GCTA stepwise conditional analysis by Mack and colleagues, adding genome-wide significant variants within 1.76 Mb of the *LPA* gene to a model based on summary statistics and LD from a reference panel, testing each SNP for association conditional on previously included SNPs. These were obtained from the Mack et al., 2017 GWAS described earlier, as opposed to the UKB GWAS used for the main genetic instruments, to avoid overfitting due to high sample overlap. The 30 SNPs prioritized by Mack et al. used as weights are presented in **Table S1**.

We then categorized the genetic score into quintiles based on its distribution. Finally, we investigated the relationship of the Lp(a) genetic score quintile with risk of APPOs by fitting a linear or logistic (according to the outcome) regression model with the Lp(a) score quintile as an unordered categorical exposure, using the fifth quintile (highest genetically predicted levels) as the reference category for comparison.

We identified and derived a weighted Lp(a) genetic score for 198,854 women of White European ancestry with linked genetic data in our UKB MR-PREG sample. Each quintile of the Lp(a) genetic score distribution contained 39,771 women.

Each SD increase in the genetic score was associated with a mean 28.85 nmol/L increase in measured Lp(a) levels (SE = 0.14, $P < 1\times10^{-308}$). The quintile groups were comparable to each other in terms of BMI and smoking rates, which can modify the risk of APPOs (**Table S10**).

Despite the Lp(a) genetic score having a normal distribution (**Figure S2**), there are very modest differences between the distribution of measured Lp(a) levels from first to fourth quintiles of the Lp(a) genetic score, with only the 5th quintile showing substantially higher levels compared to the rest (**Figure S3**).

When comparing the numbers of Lp(a) measurements not taken or not within the detectable range, a similar trend was observed (**Table S11**); the number of Lp(a) measurements not available due to being below the lower limit was comparable between the first three quartiles. Contrastingly, the 5th quintile had the largest proportion of missing measurements due to being above the upper limit and the lowest proportion of samples below the lower limit.

The number of cases for each APPO, including thrombotic events during pregnancy, across each of the genetic score quintiles is shown in **Table S12**.

The risk of each APPO (and mean difference for continuous traits) across Lp(a) genetic score quintiles, relative to the 5th quintile, is shown in **Figure S4** (**Table S13**). Association estimates for quintiles 1 through 4 were wide and frequently included the null, with no clear indication of a progressive trend or gradient across these quintiles. The only nominally significant association was an estimated lower risk of induction of labour for the 3$^{rd}$ quintile relative to the 5$^{th}$ (OR = 0.71, 95% CI= 0.60, 0.85, FDR *P* = 0.001).

#### LPA loss-of-function variants

We leveraged the use of loss-of-function (LoF) variants in the *LPA* region as a human knockout model. We identified the LoF variants rs41272114 (which was one of the genetic instruments in the MR analysis), rs41267811, rs143431368, rs200099994, rs41267813, rs41259144 and rs559580002 as reported by Coassin and colleagues and in GeneBass[13,14], and investigated how carrying two copies (homozygous) of these LoF variants related to the risk of the considered APPOs. This was evaluated by performing a linear or logistic regression with being homozygote for one of these variants as a binary exposure.

The observed MAF for each searched variant within our sample are shown in **Table S14**. Individuals missing genotype information on any of the considered variants were excluded from the analysis.

Similarly to the analysis above, the number of samples with levels outside the measurable range were compared between knockouts and the rest of the sample (**Table S15**). Most (286 out of the 292 with an attempted measurement) knockouts had levels below the detectable range. Those with detectable Lp(a) levels were still very low (**Figure S4**). On the other hand, the rest of the sample had comparable proportions of non-measured, too low and too high Lp(a) levels. The number of APPO cases according to LoF status is shown in **Table S16**.

Carriers of both copies of the same *LPA* LoF variant (from hereafter referred to as knockouts) were identified for three variants: rs41272114, rs41259144 and rs143431368. These individuals were coded as knockouts (N = 362) whilst the rest of the sample were coded as controls (N = 197,600). Further information on this analysis is presented in Section 1.3 of the supplement.

To explore the safety of low to null Lp(a) levels in pregnancy, we estimated the association between being a *LPA* knockout and the APPOs and continuous traits of interest. We did not observe robust evidence of association for any of the outcomes, although 95% confidence intervals were wide due to the relatively modest sample size (**Figure S5, Table S17**). Thrombotic events were excluded from this analysis due to having no cases in the *LPA* knockout group. As a positive control, we examined the association between being a *LPA* knockout and coronary artery disease, which would be expected to show a protective effect. This outcome was included to assess whether our study had sufficient power to detect an expected association. We observed a strong but imprecise negative association that spanned the null and did not hold up to multiple testing burden (OR = 0.53, 95% CI= 0.27, 1.03).

#### Limitations of these analyses

Lp(a) concentrations are predominantly influenced by apo(a) isoforms, and the effects of LoF variants used to weight the score in GWAS are often masked in individuals with high molecular-weight isoforms, who already have low Lp(a) levels[13]. In our GS analysis, the strong Lp(a)-lowering effect of high molecular-weight apo(a) isoforms could have outweighed the influence of variants linked to higher Lp(a), meaning some individuals were assigned scores suggesting elevated levels despite their apo(a) isoform-driven low concentrations. This may explain the modest differences in measured Lp(a) across the first to fourth genetic score quintiles. This likely explains the modest differences in measured Lp(a) across the first to fourth genetic score quintiles. Similarly, in our analysis comparing LoF individuals to the rest of the sample, the accentuated skew of Lp(a) levels towards the lower end of the distribution, likely due to the presence of other genetic determinants of low levels such as high molecular-weight isoforms, may have limited the statistical power of these comparisons.

### Supplementary Tables

**Table S1: Candidate genetic instruments for Lp(a) as derived by Shi et al., 2024, ordered according to effect size**. EA, effect allele; OA, other allele; EAF, effect allele frequency; SE, standard error. N.B 'True' refers to the SNP being present in at least one of the MR-PREG outcomes.

| **rsID** | **EA** | **OA** | **EAF** | **Effect size** | **SE** | **P-value** | **$R^2$** | **SNP or tag SNP present in MR-PREG** |
|---|---|---|---|---|---|---|---|---|
| rs151135411 | G | A | 0.9992 | -187.73 | 2.32 | 0.00E+00 | 0.0041 | False |
| rs140570886 | T | C | 0.9845 | -166.2 | 0.609 | 0.00E+00 | 0.1757 | True |
| rs41267813 | G | A | 0.9978 | 124.86 | 1.254 | 0.00E+00 | 0.0002 | False |
| rs10455872 | G | A | 0.0805 | 100.27 | 0.337 | 0.00E+00 | 0.2606 | True |
| rs147555597 | G | A | 0.9904 | -99.53 | 0.576 | 0.00E+00 | 0.0404 | True |
| rs534452768 | T | C | 0.9989 | -82.1 | 1.682 | 0.00E+00 | 0.0017 | False |
| rs4252191 | C | A | 0.9943 | -77.91 | 0.758 | 0.00E+00 | 0.0023 | True |
| rs571962116 | T | C | 0.0024 | 64.68 | 1.098 | 0.00E+00 | 0.0042 | False |
| rs2802355 | T | G | 0.0026 | 58.47 | 1.096 | 0.00E+00 | 0.0014 | False |
| rs147936725 | T | G | 0.0026 | 49.43 | 1.246 | 0.00E+00 | 0.0022 | False |
| rs142231215 | G | A | 0.9951 | -44.25 | 0.977 | 0.00E+00 | 0.0025 | False |
| rs73596816 | G | A | 0.9669 | -37.93 | 0.329 | 0.00E+00 | 0.018 | True |

| | | | | | | | | |
|---|---|---|---|---|---|---|---|---|
| rs145975837 | T | C | 0.9943 | 33.97 | 0.728 | 0.00E+00 | 0.0018 | True |
| rs191731233 | T | C | 0.0019 | 31.15 | 1.249 | 2.48E-137 | 0.0014 | True |
| rs12204009 | T | C | 0.9945 | -25.24 | 0.775 | 1.20E-232 | 0.0299 | False |
| rs41272114 | T | C | 0.0385 | -23.17 | 0.294 | 0.00E+00 | 0.011 | True |
| rs6938647 | C | A | 0.7819 | 20.6 | 0.179 | 0.00E+00 | 0.0491 | True |
| rs6905073 | T | G | 0.3428 | -17.44 | 0.164 | 0.00E+00 | 0.0242 | True |
| rs41272086 | G | A | 0.8934 | 15.81 | 0.211 | 0.00E+00 | 0.0014 | False |
| rs180785936 | G | C | 0.0048 | 12.32 | 0.848 | 8.08E-48 | 0.0152 | False |
| rs559435190 | GT | G | 0.9871 | -12.12 | 0.561 | 1.20E-103 | 0.0575 | False |
| rs550904617 | T | C | 0.0015 | 12.02 | 1.442 | 7.94E-17 | 0.0046 | False |
| rs148349043 | G | C | 0.0133 | 11.96 | 0.514 | 5.36E-120 | 0.011 | True |
| rs6932293 | T | C | 0.9839 | -11.87 | 0.483 | 3.19E-133 | 0.0168 | True |
| rs150044916 | G | A | 0.998 | -9.44 | 1.278 | 1.53E-13 | 0.0091 | False |
| rs4252198 | G | C | 0.0198 | 6.96 | 0.413 | 8.94E-64 | 0.0179 | True |
| rs9457997 | G | A | 0.8382 | -6.39 | 0.163 | 0.00E+00 | 0.0088 | True |
| rs58432601 | G | A | 0.8797 | 6.34 | 0.172 | 6.49E-297 | 0.0033 | True |

| | | | | | | | | |
|---|---|---|---|---|---|---|---|---|
| rs377175450 | AC | A | 0.0087 | 6.04 | 0.702 | 7.67E-18 | 0.0021 | True |

**Table S2:** ***Weights used in the Lp(a) genetic score*** **from Mack et al., 2017.** EAF – effect allele frequency, beta – conditional SNP association estimate, se – standard error.

| SNP | Effect allele | Other allele | EAF | beta | se | *P* value |
|---|---|---|---|---|---|---|
| rs112842440 | T | G | 0.016 | 5.4 | 1.01 | 2.04e-13 |
| rs117026595 | T | A | 0.024 | -10.07 | 1.48 | 2.61e-16 |
| rs12207325 | A | G | 0.005 | -10.49 | 1.539 | 1.53e-20 |
| rs12664092 | C | A | 0.028 | 6.35 | 0.739 | 1.85e-18 |
| rs140570886 | C | T | 0.011 | 23.78 | 1.519 | 7.49e-28 |
| rs141463285 | A | T | 0.007 | 15.42 | 2.178 | 9.42e-10 |
| rs142126734 | A | G | 0.042 | 6.09 | 0.615 | 2.42e-29 |
| rs145470851 | A | G | 0.009 | -9.76 | 1.514 | 2.34e-11 |
| rs147010904 | T | C | 0.004 | 13.04 | 1.968 | 2.93e-09 |
| rs147555597 | A | G | 0.007 | 17 | 2.453 | 7.04e-09 |
| rs149302195 | T | C | 0.005 | -7.71 | 1.328 | 2.89e-09 |
| rs182532458 | A | G | 0.006 | 6.46 | 1.199 | 3.11e-10 |
| rs186696265 | T | C | 0.011 | 24.75 | 1.755 | 4.3e-22 |
| rs188974863 | A | T | 0.093 | -5.17 | 0.496 | 2.25e-28 |

| | | | | | | |
|---|---|---|---|---|---|---|
| rs3798221 | T | G | 0.212 | -3.53 | 0.418 | 1.09e-31 |
| rs41267807 | C | T | 0.017 | -6.43 | 1.061 | 5.1e-10 |
| rs41267809 | G | A | 0.021 | -5.89 | 1.145 | 3.2e-09 |
| rs41272114 | T | C | 0.028 | -4.7 | 0.759 | 7.95e-20 |
| rs4252198 | G | C | 0.024 | 10.09 | 1.138 | 9.43e-18 |
| rs55730499 | T | C | 0.07 | 10.18 | 0.722 | 3.24e-27 |
| rs56393506 | T | C | 0.171 | 3.12 | 0.445 | 7.77e-13 |
| rs59614420 | A | G | 0.279 | -2.11 | 0.289 | 5.85e-18 |
| rs62440901 | T | C | 0.148 | 2.6 | 0.348 | 3.92e-16 |
| rs6938647 | A | C | 0.219 | 4.12 | 0.377 | 5.75e-43 |
| rs75234242 | A | G | 0.037 | -6.49 | 0.802 | 1.36e-20 |
| rs75692336 | A | C | 0.142 | -7.38 | 0.469 | 4.91e-85 |
| rs7769879 | C | G | 0.358 | 4.69 | 0.373 | 1.05e-35 |
| rs78439586 | A | G | 0.063 | -3.34 | 0.589 | 2.32e-08 |
| rs9295143 | G | C | 0.047 | -3.09 | 0.563 | 3.74e-09 |
| rs9365169 | G | C | 0.488 | -2.91 | 0.36 | 2.53e-13 |

**Table S3: Mendelian randomization estimates for the estimated effect of Lp(a) lowering on adverse pregnancy and perinatal outcomes.** $^{-1}$ denotes MR estimates reflecting the effect of lowering Lp(a) by 1 nmol/L, $^{-210}$ denotes MR estimates reflecting the effect of lowering Lp(a) by 210 nmol/L. OR, odds ratio (binary outcome); beta, mean difference (continuous outcome); SE, standard error of beta/log odds estimate; UCI, upper 95% confidence interval; LCI, lower 95% confidence interval; FDR, False discovery rate.

| **Outcome** | **nsnp** | **F** | **$r^2$** | **OR/beta$^{(-1)}$** | **SE$^{(-1)}$** | **UCI$^{(-1)}$** | **LCI$^{(-1)}$** | **OR/beta$^{(-210)}$** | **UCI$^{(-210)}$** | **LCI$^{(-210)}$** | **P-value** | **FDR P-value** |
|---|---|---|---|---|---|---|---|---|---|---|---|---|
| Hypertensive disorders of pregnancy | 11 | 18188.5 | 0.58 | 1.0001 | 0.0001 | 1.0004 | 1.0004 | 1.021 | 1.082 | 0.964 | 0.48 | 0.89 |
| Gestational hypertension | 11 | 18188.5 | 0.58 | 1.0000 | 0.0002 | 1.0005 | 1.0005 | 1.008 | 1.110 | 0.915 | 0.88 | 0.97 |
| Preeclampsia | 12 | 16835.8 | 0.60 | 1.0001 | 0.0003 | 1.0007 | 1.0007 | 1.030 | 1.150 | 0.923 | 0.59 | 0.89 |
| Gestational diabetes | 11 | 18188.5 | 0.58 | 0.9999 | 0.0002 | 1.0004 | 1.0004 | 0.975 | 1.078 | 0.882 | 0.62 | 0.89 |
| Miscarriage | 13 | 17612.8 | 0.64 | 1.0000 | 0.0001 | 1.0002 | 1.0002 | 0.999 | 1.049 | 0.952 | 0.97 | 0.99 |
| Recurrent miscarriage | 13 | 17612.8 | 0.64 | 1.0003 | 0.0003 | 1.0009 | 1.0009 | 1.066 | 1.208 | 0.941 | 0.31 | 0.70 |
| Stillbirth | 13 | 17612.8 | 0.64 | 1.0004 | 0.0002 | 1.0008 | 1.0008 | 1.091 | 1.188 | 1.002 | 0.05 | 0.27 |
| Induction of labour | 12 | 18057.3 | 0.60 | 1.0000 | 0.0002 | 1.0003 | 1.0003 | 0.990 | 1.072 | 0.914 | 0.81 | 0.95 |
| Pre-labour rupture of membrane | 12 | 18057.3 | 0.60 | 1.0003 | 0.0002 | 1.0006 | 1.0006 | 1.063 | 1.138 | 0.993 | 0.08 | 0.27 |
| Caesarian section | 11 | 18188.5 | 0.58 | 1.0003 | 0.0002 | 1.0006 | 1.0006 | 1.054 | 1.136 | 0.978 | 0.17 | 0.48 |
| Spontaneous pre-term birth | 12 | 17481.9 | 0.60 | 0.9997 | 0.0002 | 1.0000 | 1.0000 | 0.933 | 1.005 | 0.865 | 0.07 | 0.27 |
| Small for gestational age | 12 | 18057.3 | 0.60 | 1.0001 | 0.0004 | 1.0009 | 1.0009 | 1.032 | 1.218 | 0.874 | 0.71 | 0.89 |
| Large for gestational age | 12 | 18057.3 | 0.60 | 0.9999 | 0.0002 | 1.0004 | 1.0004 | 0.981 | 1.081 | 0.889 | 0.69 | 0.89 |

| | | | | | | | | | | | | |
|---|---|---|---|---|---|---|---|---|---|---|---|---|
| Low Apgar score at 1 minute | 12 | 18057.3 | 0.60 | 1.0005 | 0.0003 | 1.0010 | 1.0010 | 1.107 | 1.235 | 0.993 | 0.07 | 0.27 |
| Low Apgar score at 5 minutes | 12 | 18057.3 | 0.60 | 1.0004 | 0.0007 | 1.0017 | 1.0017 | 1.081 | 1.440 | 0.812 | 0.59 | 0.89 |
| Postpartum haemorrhage | 13 | 11281.3 | 0.71 | 1.0000 | 0.0002 | 1.0004 | 1.0004 | 1.001 | 1.077 | 0.929 | 0.99 | 0.99 |
| NICU admission | 11 | 18501.3 | 0.55 | 0.9998 | 0.0002 | 1.0003 | 1.0003 | 0.966 | 1.068 | 0.874 | 0.50 | 0.89 |
| Congenital malformations (any) | 12 | 18057.3 | 0.60 | 0.9991 | 0.0003 | 0.9997 | 0.9997 | 0.821 | 0.939 | 0.719 | 0.00 | 0.08 |
| Gestational age | 12 | 17481.9 | 0.60 | 0.0002 | 0.0001 | 0.000 | 0.0000 | 0.0000 | 0.070 | 0.009 | 0.01 | 0.12 |
| Birth weight | 13 | 17612.8 | 0.64 | 0.0000 | 0.0000 | 0.000 | 0.0001 | 0.0000 | 0.021 | -0.005 | 0.25 | 0.63 |

**Table S4: Mendelian randomization estimates for the estimated effect of Lp(a) lowering on adverse pregnancy and perinatal outcomes using alternative exposure datasets.** $^{-1}$ denotes MR estimates reflecting the effect of lowering Lp(a) by 1 nmol/L, $^{-210}$ denotes MR estimates reflecting the effect of lowering Lp(a) by 210 nmol/L. OR, odds ratio (binary outcome); beta, mean difference (continuous outcome); SE, standard error of beta/log odds estimate; UCI, upper 95% confidence interval; LCI, lower 95% confidence interval; FDR, False discovery rate.

| Outcome | Exposure dataset | nsnp | F | OR/beta$^{(-1)}$ | SE$^{(-1)}$ | UCI$^{(-1)}$ | LCI$^{(-1)}$ | OR/beta$^{(-210)}$ | UCI$^{(-210)}$ | LCI$^{(-210)}$ | P-value | FDR P-value |
|---|---|---|---|---|---|---|---|---|---|---|---|---|
| Hypertensive disorders of pregnancy | Lp(a), apo(a) isoform-adjusted (mg/dL) | 19 | 313.4 | 1.0002 | 0.0004 | 1.0010 | 0.9995 | 1.023 | 1.102 | 0.950 | 0.551 | 0.721 |
| | LPA, Olink UKB-PPP | 24 | 370.0 | 1.0116 | 0.0122 | 1.0362 | 0.9876 | 1.012 | 1.036 | 0.988 | 0.346 | 0.557 |
| Gestational hypertension | Lp(a), apo(a) isoform-adjusted (mg/dL) | 19 | 313.4 | 1.0001 | 0.0005 | 1.0011 | 0.9992 | 1.011 | 1.110 | 0.921 | 0.815 | 0.931 |
| | LPA, Olink UKB-PPP | 24 | 370.0 | 1.0025 | 0.0151 | 1.0327 | 0.9733 | 1.003 | 1.033 | 0.973 | 0.867 | 0.947 |
| Preeclampsia | Lp(a), apo(a) isoform-adjusted (mg/dL) | 22 | 369.8 | 1.0010 | 0.0005 | 1.0020 | 1.0000 | 1.100 | 1.215 | 0.996 | 0.060 | 0.149 |
| | LPA, Olink UKB-PPP | 23 | 367.9 | 1.0607 | 0.0198 | 1.1027 | 1.0203 | 1.061 | 1.103 | 1.020 | 0.003 | 0.015 |
| Gestational diabetes | Lp(a), apo(a) isoform-adjusted (mg/dL) | 19 | 313.4 | 0.9991 | 0.0006 | 1.0003 | 0.9979 | 0.916 | 1.031 | 0.813 | 0.147 | 0.294 |
| | LPA, Olink UKB-PPP | 24 | 370.0 | 1.0003 | 0.0160 | 1.0321 | 0.9695 | 1.000 | 1.032 | 0.969 | 0.984 | 0.984 |
| Miscarriage | Lp(a), apo(a) isoform-adjusted (mg/dL) | 24 | 375.4 | 0.9995 | 0.0002 | 0.9999 | 0.9992 | 0.956 | 0.990 | 0.923 | 0.012 | 0.046 |
| | LPA, Olink UKB-PPP | 36 | 310.7 | 1.0042 | 0.0072 | 1.0185 | 0.9902 | 1.004 | 1.018 | 0.990 | 0.559 | 0.721 |

| | | | | | | | | | | | | |
|---|---|---|---|---|---|---|---|---|---|---|---|---|
| Recurrent miscarriage | Lp(a), apo(a) isoform-adjusted (mg/dL) | 24 | 375.4 | 0.9999 | 0.0006 | 1.0012 | 0.9987 | 0.995 | 1.123 | 0.881 | 0.933 | 0.957 |
| | LPA, Olink UKB-PPP | 36 | 310.7 | 0.9822 | 0.0290 | 1.0396 | 0.9279 | 0.982 | 1.040 | 0.928 | 0.535 | 0.721 |
| Stillbirth | Lp(a), apo(a) isoform-adjusted (mg/dL) | 24 | 375.4 | 1.0020 | 0.0005 | 1.0030 | 1.0010 | 1.220 | 1.345 | 1.106 | 0.000 | 0.001 |
| | LPA, Olink UKB-PPP | 36 | 310.7 | 1.0167 | 0.0237 | 1.0651 | 0.9705 | 1.017 | 1.065 | 0.970 | 0.485 | 0.719 |
| Induction of labour | Lp(a), apo(a) isoform-adjusted (mg/dL) | 19 | 385.2 | 0.9983 | 0.0009 | 1.0000 | 0.9965 | 0.843 | 1.002 | 0.709 | 0.053 | 0.142 |
| | LPA, Olink UKB-PPP | 20 | 410.8 | 0.9446 | 0.0219 | 0.9859 | 0.9050 | 0.945 | 0.986 | 0.905 | 0.009 | 0.040 |
| Pre-labour rupture of membrane | Lp(a), apo(a) isoform-adjusted (mg/dL) | 22 | 386.1 | 1.0009 | 0.0005 | 1.0018 | 0.9999 | 1.089 | 1.195 | 0.993 | 0.071 | 0.161 |
| | LPA, Olink UKB-PPP | 24 | 378.5 | 1.0137 | 0.0149 | 1.0438 | 0.9845 | 1.014 | 1.044 | 0.984 | 0.362 | 0.557 |
| Caesarian section | Lp(a), apo(a) isoform-adjusted (mg/dL) | 19 | 313.4 | 1.0013 | 0.0004 | 1.0021 | 1.0005 | 1.131 | 1.224 | 1.046 | 0.002 | 0.012 |
| | LPA, Olink UKB-PPP | 24 | 370.0 | 1.0148 | 0.0149 | 1.0448 | 0.9857 | 1.015 | 1.045 | 0.986 | 0.322 | 0.557 |
| Spontaneous pre-term birth | Lp(a), apo(a) isoform-adjusted (mg/dL) | 22 | 369.8 | 0.9988 | 0.0004 | 0.9996 | 0.9981 | 0.890 | 0.958 | 0.827 | 0.002 | 0.012 |
| | LPA, Olink UKB-PPP | 25 | 372.9 | 0.9630 | 0.0163 | 0.9942 | 0.9327 | 0.963 | 0.994 | 0.933 | 0.021 | 0.073 |

| | | | | | | | | | | | | |
|---|---|---|---|---|---|---|---|---|---|---|---|---|
| Small for gestational age | Lp(a), apo(a) isoform-adjusted (mg/dL) | 19 | 385.2 | 0.9999 | 0.0011 | 1.0020 | 0.9978 | 0.989 | 1.211 | 0.808 | 0.917 | 0.957 |
| | LPA, Olink UKB-PPP | 20 | 383.2 | 1.0080 | 0.0315 | 1.0722 | 0.9476 | 1.008 | 1.072 | 0.948 | 0.801 | 0.931 |
| Large for gestational age | Lp(a), apo(a) isoform-adjusted (mg/dL) | 18 | 301.7 | 1.0013 | 0.0008 | 1.0030 | 0.9997 | 1.139 | 1.336 | 0.970 | 0.112 | 0.236 |
| | LPA, Olink UKB-PPP | 21 | 399.9 | 1.0331 | 0.0241 | 1.0830 | 0.9855 | 1.033 | 1.083 | 0.986 | 0.175 | 0.327 |
| Low Apgar score at 1 minute | Lp(a), apo(a) isoform-adjusted (mg/dL) | 18 | 406.2 | 1.0015 | 0.0009 | 1.0032 | 0.9999 | 1.163 | 1.371 | 0.986 | 0.073 | 0.161 |
| | LPA, Olink UKB-PPP | 20 | 409.7 | 1.0050 | 0.0320 | 1.0701 | 0.9439 | 1.005 | 1.070 | 0.944 | 0.876 | 0.947 |
| Low Apgar score at 5 minutes | Lp(a), apo(a) isoform-adjusted (mg/dL) | 19 | 385.2 | 1.0013 | 0.0030 | 1.0072 | 0.9955 | 1.138 | 2.014 | 0.642 | 0.658 | 0.823 |
| | LPA, Olink UKB-PPP | 21 | 399.9 | 1.0453 | 0.0728 | 1.2057 | 0.9062 | 1.045 | 1.206 | 0.906 | 0.543 | 0.721 |
| Postpartum haemorrhage | Lp(a), apo(a) isoform-adjusted (mg/dL) | 24 | 319.7 | 0.9999 | 0.0004 | 1.0007 | 0.9990 | 0.986 | 1.073 | 0.907 | 0.744 | 0.902 |
| | LPA, Olink UKB-PPP | 36 | 315.8 | 0.9836 | 0.0179 | 1.0187 | 0.9496 | 0.984 | 1.019 | 0.950 | 0.355 | 0.557 |
| NICU admission | Lp(a), apo(a) isoform-adjusted (mg/dL) | 18 | 388.1 | 0.9975 | 0.0008 | 0.9990 | 0.9959 | 0.782 | 0.910 | 0.672 | 0.001 | 0.012 |
| | LPA, Olink UKB-PPP | 21 | 399.9 | 0.9333 | 0.0316 | 0.9929 | 0.8773 | 0.933 | 0.993 | 0.877 | 0.029 | 0.088 |

| | | | | | | | | | | | | |
|---|---|---|---|---|---|---|---|---|---|---|---|---|
| Congenital malformations (any) | Lp(a), apo(a) isoform-adjusted (mg/dL) | 19 | 385.2 | 0.9966 | 0.0011 | 0.9987 | 0.9945 | 0.717 | 0.879 | 0.584 | 0.001 | 0.012 |
| | LPA, Olink UKB-PPP | 21 | 399.9 | 0.9530 | 0.0359 | 1.0225 | 0.8883 | 0.953 | 1.022 | 0.888 | 0.180 | 0.327 |
| Gestational age | Lp(a), apo(a) isoform-adjusted (mg/dL) | 22 | 374.6 | 0.0009 | 0.0002 | 0.0013 | 0.0006 | 0.092 | 0.128 | 0.056 | 0.000 | 0.000 |
| | LPA, Olink UKB-PPP | 28 | 350.6 | 0.0193 | 0.0084 | 0.0359 | 0.0028 | 0.019 | 0.036 | 0.003 | 0.022 | 0.073 |
| Birth weight | Lp(a), apo(a) isoform-adjusted (mg/dL) | 24 | 375.4 | 0.0002 | 8.06906572891004e-05 | 0.0004 | 0.0001 | 0.024 | 0.040 | 0.009 | 0.002 | 0.012 |
| | LPA, Olink UKB-PPP | 36 | 310.7 | 0.0077 | 0.0037 | 0.0150 | 0.0004 | 0.008 | 0.015 | 0.000 | 0.037 | 0.107 |

**Table S5: Mendelian randomization estimates for the estimated effect of Lp(a) lowering on adverse pregnancy and perinatal outcomes, unadjusted and adjusted for fetal genotype.** $^{-1}$ denotes MR estimates reflecting the effect of lowering Lp(a) by 1 nmol/L, $^{-210}$ denotes MR estimates reflecting the effect of lowering Lp(a) by 210 nmol/L. OR, odds ratio (binary outcome); beta, mean difference (continuous outcome); SE, standard error of beta/log odds estimate; UCI, upper 95% confidence interval; LCI, lower 95% confidence interval; FDR, False discovery rate.

| **Outcome** | **SNP-outcome estimates used** | **nsnp** | **OR/beta$^{(-1)}$** | **SE$^{(-1)}$** | **UCI$^{(-1)}$** | **LCI$^{(-1)}$** | **OR/beta$^{(-210)}$** | **UCI$^{(-210)}$** | **LCI$^{(-210)}$** | **P-value** | **FDR P-value** |
|---|---|---|---|---|---|---|---|---|---|---|---|
| Hypertensive disorders of pregnancy | Maternal (unadjusted) | 10 | 1.0001 | 0.0001 | 1.0003 | 0.9998 | 1.011 | 1.069 | 0.956 | 0.70 | 0.97 |
| | Maternal (fetal adjusted) | 10 | 1.0001 | 0.0002 | 1.0005 | 0.9997 | 1.016 | 1.109 | 0.931 | 0.72 | 0.97 |
| Gestational hypertension | Maternal (unadjusted) | 11 | 1.0000 | 0.0002 | 1.0005 | 0.9996 | 1.008 | 1.107 | 0.919 | 0.86 | 0.97 |
| | Maternal (fetal adjusted) | 11 | 1.0001 | 0.0003 | 1.0006 | 0.9996 | 1.012 | 1.123 | 0.912 | 0.82 | 0.97 |

| | | | | | | | | | | | |
|---|---|---|---|---|---|---|---|---|---|---|---|
| Preeclampsia | Maternal (unadjusted) | 11 | 1.0002 | 0.0003 | 1.0008 | 0.9997 | 1.047 | 1.171 | 0.936 | 0.42 | 0.97 |
| | Maternal (fetal adjusted) | 11 | 1.0004 | 0.0004 | 1.0011 | 0.9997 | 1.084 | 1.256 | 0.937 | 0.28 | 0.94 |
| Gestational diabetes | Maternal (unadjusted) | 9 | 0.9995 | 0.0002 | 1.0000 | 0.9991 | 0.904 | 0.997 | 0.820 | 0.04 | 0.36 |
| | Maternal (fetal adjusted) | 9 | 0.9999 | 0.0006 | 1.0010 | 0.9988 | 0.980 | 1.241 | 0.773 | 0.87 | 0.97 |
| Induction of labour | Maternal (unadjusted) | 12 | 1.0000 | 0.0002 | 1.0003 | 0.9996 | 0.990 | 1.072 | 0.914 | 0.81 | 0.97 |
| | Maternal (fetal adjusted) | 12 | 0.9999 | 0.0002 | 1.0003 | 0.9994 | 0.972 | 1.070 | 0.882 | 0.56 | 0.97 |
| Pre-labour rupture of membrane | Maternal (unadjusted) | 11 | 1.0003 | 0.0002 | 1.0006 | 1.0000 | 1.073 | 1.144 | 1.006 | 0.03 | 0.30 |
| | Maternal (fetal adjusted) | 11 | 1.0005 | 0.0002 | 1.0010 | 1.0001 | 1.113 | 1.221 | 1.015 | 0.02 | 0.24 |
| Caesarian section | Maternal (unadjusted) | 11 | 1.0002 | 0.0002 | 1.0006 | 0.9999 | 1.053 | 1.139 | 0.973 | 0.20 | 0.90 |
| | Maternal (fetal adjusted) | 11 | 1.0003 | 0.0003 | 1.0010 | 0.9997 | 1.070 | 1.224 | 0.936 | 0.32 | 0.94 |
| Spontaneous pre-term birth | Maternal (unadjusted) | 12 | 0.9997 | 0.0002 | 1.0000 | 0.9994 | 0.939 | 1.005 | 0.878 | 0.07 | 0.40 |
| | Maternal (fetal adjusted) | 12 | 0.9996 | 0.0004 | 1.0003 | 0.9989 | 0.919 | 1.064 | 0.794 | 0.26 | 0.94 |
| Small for gestational age | Maternal (unadjusted) | 11 | 1.0002 | 0.0004 | 1.0010 | 0.9994 | 1.036 | 1.221 | 0.879 | 0.67 | 0.97 |
| | Maternal (fetal adjusted) | 11 | 1.0000 | 0.0004 | 1.0008 | 0.9992 | 0.995 | 1.180 | 0.840 | 0.96 | 0.98 |
| Large for gestational age | Maternal (unadjusted) | 12 | 0.9999 | 0.0002 | 1.0004 | 0.9994 | 0.981 | 1.081 | 0.889 | 0.69 | 0.97 |
| | Maternal (fetal adjusted) | 12 | 1.0000 | 0.0004 | 1.0008 | 0.9992 | 1.002 | 1.186 | 0.847 | 0.98 | 0.98 |
| Low Apgar score at 1 minute | Maternal (unadjusted) | 12 | 1.0005 | 0.0003 | 1.0010 | 1.0000 | 1.107 | 1.235 | 0.993 | 0.07 | 0.40 |
| | Maternal (fetal adjusted) | 12 | 1.0006 | 0.0003 | 1.0012 | 0.9999 | 1.125 | 1.295 | 0.977 | 0.10 | 0.52 |
| Low Apgar score at 5 minutes | Maternal (unadjusted) | 12 | 1.0004 | 0.0007 | 1.0017 | 0.9990 | 1.081 | 1.440 | 0.812 | 0.59 | 0.97 |
| | Maternal (fetal adjusted) | 12 | 1.0005 | 0.0010 | 1.0025 | 0.9985 | 1.107 | 1.683 | 0.727 | 0.64 | 0.97 |

| | | | | | | | | | | | |
|---|---|---|---|---|---|---|---|---|---|---|---|
| NICU admission | Maternal (unadjusted) | 11 | 0.9998 | 0.0002 | 1.0003 | 0.9994 | 0.966 | 1.068 | 0.874 | 0.50 | 0.97 |
| | Maternal (fetal adjusted) | 11 | 0.9997 | 0.0003 | 1.0003 | 0.9991 | 0.942 | 1.070 | 0.829 | 0.36 | 0.94 |
| Congenital malformations (any) | Maternal (unadjusted) | 10 | 0.9991 | 0.0003 | 0.9996 | 0.9986 | 0.827 | 0.924 | 0.740 | 0.00 | 0.07 |
| | Maternal (fetal adjusted) | 10 | 0.9990 | 0.0003 | 0.9997 | 0.9983 | 0.815 | 0.941 | 0.707 | 0.01 | 0.17 |
| Gestational age | Maternal (unadjusted) | 12 | 0.0002 | 0.0001 | 0.0003 | 0.0000 | 0.039 | 0.070 | 0.009 | 0.01 | 0.17 |
| | Maternal (fetal adjusted) | 12 | 0.0002 | 0.0001 | 0.0004 | 0.0000 | 0.042 | 0.085 | -0.001 | 0.06 | 0.38 |
| Birth weight | Maternal (unadjusted) | 13 | 0.0000 | 0.0000 | 0.0001 | 0.0000 | 0.008 | 0.021 | -0.005 | 0.25 | 0.94 |
| | Maternal (fetal adjusted) | 13 | 0.0001 | 0.0000 | 0.0001 | 0.0000 | 0.011 | 0.029 | -0.007 | 0.24 | 0.94 |

**Table S6: Leave-one-study-out Mendelian randomization estimates for the estimated effect of Lp(a) lowering on adverse pregnancy and perinatal outcomes.** $^{-1}$ denotes MR estimates reflecting the effect of lowering Lp(a) by 1 nmol/L, $^{-210}$ denotes MR estimates reflecting the effect of lowering Lp(a) by 210 nmol/L. OR, odds ratio (binary outcome); beta, mean difference (continuous outcome); SE, standard error of beta/log odds estimate; UCI, upper 95% confidence interval; LCI, lower 95% confidence interval; FDR, False discovery rate.

| Outcome | Study removed | OR/beta$^{(-1)}$ | SE$^{(-1)}$ | UCI$^{(-1)}$ | LCI$^{(-1)}$ | OR/beta$^{(-210)}$ | UCI$^{(-210)}$ | LCI$^{(-210)}$ | P-value | FDR P-value |
|---|---|---|---|---|---|---|---|---|---|---|
| Hypertensive disorders of pregnancy | -ALSPAC | 1.0001 | 0.0002 | 1.0004 | 0.9998 | 1.021 | 1.093 | 0.953 | 0.56 | 0.86 |
| | -BIB-SA | 1.0001 | 0.0002 | 1.0004 | 0.9998 | 1.021 | 1.091 | 0.955 | 0.55 | 0.86 |
| | -BIB-WE | 1.0001 | 0.0002 | 1.0004 | 0.9998 | 1.017 | 1.088 | 0.951 | 0.62 | 0.89 |
| | -FinnGen | 1.0001 | 0.0002 | 1.0005 | 0.9997 | 1.030 | 1.120 | 0.947 | 0.49 | 0.84 |
| | -MOBA | 1.0000 | 0.0002 | 1.0005 | 0.9996 | 1.010 | 1.111 | 0.918 | 0.84 | 0.96 |
| | -UKB | 1.0001 | 0.0002 | 1.0004 | 0.9998 | 1.025 | 1.097 | 0.959 | 0.46 | 0.84 |
| Gestational hypertension | -ALSPAC | 1.0001 | 0.0002 | 1.0005 | 0.9996 | 1.015 | 1.118 | 0.921 | 0.76 | 0.91 |
| | -BIB-SA | 1.0001 | 0.0002 | 1.0005 | 0.9997 | 1.020 | 1.119 | 0.930 | 0.68 | 0.89 |
| | -BIB-WE | 1.0001 | 0.0002 | 1.0005 | 0.9996 | 1.012 | 1.113 | 0.920 | 0.80 | 0.93 |
| | -FinnGen | 1.0002 | 0.0003 | 1.0007 | 0.9997 | 1.038 | 1.153 | 0.933 | 0.49 | 0.84 |
| | -MOBA | 1.0001 | 0.0004 | 1.0008 | 0.9993 | 1.011 | 1.174 | 0.871 | 0.88 | 0.97 |
| | -UKB | 1.0001 | 0.0002 | 1.0006 | 0.9997 | 1.024 | 1.125 | 0.932 | 0.62 | 0.89 |
| Preeclampsia | -BIB-SA | 1.0001 | 0.0002 | 1.0005 | 0.9996 | 1.014 | 1.108 | 0.929 | 0.75 | 0.91 |
| | -BIB-WE | 1.0001 | 0.0002 | 1.0005 | 0.9997 | 1.016 | 1.110 | 0.930 | 0.73 | 0.91 |
| | -FinnGen | 1.0000 | 0.0003 | 1.0006 | 0.9994 | 0.999 | 1.129 | 0.884 | 0.99 | 0.99 |
| | -MOBA | 1.0001 | 0.0002 | 1.0005 | 0.9996 | 1.014 | 1.112 | 0.923 | 0.78 | 0.91 |

| | | | | | | | | | | |
|---|---|---|---|---|---|---|---|---|---|---|
| | -Public | 1.0001 | 0.0003 | 1.0006 | 0.9996 | 1.020 | 1.137 | 0.915 | 0.72 | 0.91 |
| | -UKB | 1.0001 | 0.0002 | 1.0006 | 0.9997 | 1.025 | 1.123 | 0.935 | 0.60 | 0.89 |
| Gestational diabetes | -FinnGen | 0.9996 | 0.0004 | 1.0003 | 0.9989 | 0.917 | 1.060 | 0.794 | 0.24 | 0.70 |
| | -MOBA | 0.9998 | 0.0002 | 1.0003 | 0.9993 | 0.952 | 1.054 | 0.860 | 0.34 | 0.74 |
| | -Public | 1.0000 | 0.0003 | 1.0006 | 0.9995 | 1.004 | 1.130 | 0.893 | 0.94 | 0.99 |
| Miscarriage | -ALSPAC | 0.9999 | 0.0001 | 1.0001 | 0.9998 | 0.985 | 1.017 | 0.954 | 0.35 | 0.74 |
| | -BIB-SA | 0.9999 | 0.0001 | 1.0001 | 0.9998 | 0.983 | 1.014 | 0.952 | 0.28 | 0.72 |
| | -BIB-WE | 0.9999 | 0.0001 | 1.0001 | 0.9998 | 0.983 | 1.015 | 0.953 | 0.30 | 0.72 |
| | -FinnGen | 0.9999 | 0.0001 | 1.0001 | 0.9997 | 0.978 | 1.011 | 0.946 | 0.19 | 0.67 |
| | -MOBA | 0.9999 | 0.0001 | 1.0001 | 0.9998 | 0.981 | 1.014 | 0.949 | 0.26 | 0.72 |
| | -UKB | 1.0000 | 0.0002 | 1.0003 | 0.9997 | 1.000 | 1.070 | 0.935 | 0.99 | 0.99 |
| Recurrent miscarriage | -ALSPAC | 1.0003 | 0.0003 | 1.0009 | 0.9997 | 1.059 | 1.202 | 0.933 | 0.37 | 0.74 |
| | -FinnGen | 1.0003 | 0.0003 | 1.0009 | 0.9996 | 1.062 | 1.213 | 0.929 | 0.38 | 0.74 |
| | -MOBA | 1.0001 | 0.0003 | 1.0008 | 0.9994 | 1.021 | 1.171 | 0.891 | 0.76 | 0.91 |
| | -UKB | 1.0003 | 0.0005 | 1.0013 | 0.9993 | 1.064 | 1.321 | 0.856 | 0.58 | 0.88 |
| Stillbirth | -ALSPAC | 1.0004 | 0.0002 | 1.0008 | 1.0000 | 1.096 | 1.194 | 1.007 | 0.03 | 0.33 |
| | -BIB-SA | 1.0004 | 0.0002 | 1.0008 | 1.0000 | 1.095 | 1.193 | 1.006 | 0.04 | 0.33 |
| | -BIB-WE | 1.0004 | 0.0002 | 1.0008 | 1.0000 | 1.097 | 1.195 | 1.008 | 0.03 | 0.33 |
| | -MOBA | 1.0004 | 0.0002 | 1.0008 | 1.0000 | 1.094 | 1.192 | 1.004 | 0.04 | 0.33 |
| | -UKB | 0.9996 | 0.0011 | 1.0019 | 0.9974 | 0.926 | 1.478 | 0.581 | 0.75 | 0.91 |

| | | | | | | | | | | |
|---|---|---|---|---|---|---|---|---|---|---|
| Induction of labour | -ALSPAC | 0.9998 | 0.0002 | 1.0001 | 0.9995 | 0.958 | 1.027 | 0.894 | 0.22 | 0.70 |
| | -BIB-SA | 0.9999 | 0.0002 | 1.0002 | 0.9996 | 0.978 | 1.046 | 0.914 | 0.52 | 0.86 |
| | -BIB-WE | 1.0000 | 0.0002 | 1.0003 | 0.9996 | 0.994 | 1.065 | 0.928 | 0.86 | 0.97 |
| | -MOBA | 0.9994 | 0.0003 | 1.0001 | 0.9987 | 0.886 | 1.023 | 0.768 | 0.10 | 0.54 |
| | -UKB | 1.0000 | 0.0002 | 1.0003 | 0.9997 | 1.001 | 1.074 | 0.933 | 0.98 | 0.99 |
| Pre-labour rupture of membrane | -ALSPAC | 1.0002 | 0.0002 | 1.0006 | 0.9999 | 1.047 | 1.130 | 0.970 | 0.24 | 0.70 |
| | -FinnGen | 1.0001 | 0.0002 | 1.0006 | 0.9996 | 1.021 | 1.123 | 0.928 | 0.67 | 0.89 |
| | -MOBA | 1.0005 | 0.0003 | 1.0010 | 1.0000 | 1.115 | 1.241 | 1.001 | 0.05 | 0.35 |
| Caesarian section | -ALSPAC | 1.0002 | 0.0001 | 1.0005 | 0.9999 | 1.043 | 1.102 | 0.987 | 0.14 | 0.58 |
| | -BIB-SA | 1.0002 | 0.0001 | 1.0005 | 0.9999 | 1.044 | 1.103 | 0.988 | 0.12 | 0.58 |
| | -BIB-WE | 1.0002 | 0.0001 | 1.0005 | 0.9999 | 1.043 | 1.102 | 0.986 | 0.14 | 0.58 |
| | -FinnGen | 1.0002 | 0.0002 | 1.0005 | 0.9999 | 1.052 | 1.120 | 0.989 | 0.11 | 0.55 |
| | -MOBA | 1.0004 | 0.0002 | 1.0009 | 1.0000 | 1.093 | 1.196 | 0.999 | 0.05 | 0.37 |
| | -UKB | 1.0002 | 0.0001 | 1.0005 | 0.9999 | 1.044 | 1.105 | 0.986 | 0.14 | 0.58 |
| Spontaneous pre-term birth | -BIB-SA | 0.9997 | 0.0001 | 1.0000 | 0.9994 | 0.938 | 0.995 | 0.884 | 0.03 | 0.33 |
| | -BIB-WE | 0.9997 | 0.0001 | 1.0000 | 0.9994 | 0.939 | 0.997 | 0.885 | 0.04 | 0.33 |
| | -MOBA | 0.9997 | 0.0002 | 1.0000 | 0.9994 | 0.945 | 1.007 | 0.886 | 0.08 | 0.51 |
| | -Public | 0.9996 | 0.0004 | 1.0003 | 0.9989 | 0.924 | 1.069 | 0.800 | 0.29 | 0.72 |
| | -UKB | 0.9997 | 0.0001 | 0.9999 | 0.9994 | 0.929 | 0.986 | 0.876 | 0.02 | 0.33 |
| Small for gestational age | -ALSPAC | 1.0002 | 0.0004 | 1.0009 | 0.9995 | 1.047 | 1.210 | 0.906 | 0.53 | 0.86 |

| | | | | | | | | | | |
|---|---|---|---|---|---|---|---|---|---|---|
| | -BIB-SA | 1.0004 | 0.0003 | 1.0011 | 0.9997 | 1.092 | 1.261 | 0.946 | 0.23 | 0.70 |
| | -BIB-WE | 1.0001 | 0.0004 | 1.0008 | 0.9994 | 1.023 | 1.187 | 0.881 | 0.77 | 0.91 |
| | -MOBA | 1.0006 | 0.0005 | 1.0017 | 0.9996 | 1.144 | 1.422 | 0.920 | 0.23 | 0.70 |
| | -UKB | 1.0003 | 0.0004 | 1.0010 | 0.9996 | 1.067 | 1.243 | 0.915 | 0.41 | 0.78 |
| Large for gestational age | -ALSPAC | 1.0000 | 0.0002 | 1.0004 | 0.9995 | 0.995 | 1.090 | 0.908 | 0.91 | 0.99 |
| | -BIB-SA | 1.0000 | 0.0002 | 1.0004 | 0.9996 | 0.997 | 1.090 | 0.912 | 0.94 | 0.99 |
| | -BIB-WE | 1.0000 | 0.0002 | 1.0004 | 0.9995 | 0.993 | 1.087 | 0.907 | 0.88 | 0.97 |
| | -MOBA | 0.9997 | 0.0004 | 1.0005 | 0.9988 | 0.937 | 1.119 | 0.784 | 0.47 | 0.84 |
| | -UKB | 1.0000 | 0.0002 | 1.0005 | 0.9995 | 0.999 | 1.101 | 0.907 | 0.98 | 0.99 |
| Low Apgar score at 1 minute | -ALSPAC | 1.0004 | 0.0003 | 1.0009 | 0.9998 | 1.082 | 1.211 | 0.966 | 0.18 | 0.65 |
| | -BIB-SA | 1.0004 | 0.0003 | 1.0010 | 0.9999 | 1.099 | 1.224 | 0.986 | 0.09 | 0.51 |
| | -BIB-WE | 1.0006 | 0.0003 | 1.0011 | 1.0000 | 1.123 | 1.254 | 1.005 | 0.04 | 0.33 |
| | -MOBA | 1.0006 | 0.0006 | 1.0018 | 0.9995 | 1.141 | 1.461 | 0.891 | 0.29 | 0.72 |
| Low Apgar score at 5 minutes | -BIB-SA | 1.0003 | 0.0007 | 1.0016 | 0.9990 | 1.060 | 1.394 | 0.806 | 0.68 | 0.89 |
| | -BIB-WE | 1.0003 | 0.0007 | 1.0017 | 0.9990 | 1.069 | 1.419 | 0.805 | 0.64 | 0.89 |
| | -MOBA | 0.9991 | 0.0020 | 1.0030 | 0.9951 | 0.820 | 1.873 | 0.359 | 0.64 | 0.89 |
| NICU admission | -ALSPAC | 0.9998 | 0.0003 | 1.0003 | 0.9992 | 0.956 | 1.070 | 0.854 | 0.43 | 0.80 |
| | -MOBA | 1.0010 | 0.0009 | 1.0027 | 0.9992 | 1.226 | 1.774 | 0.848 | 0.28 | 0.72 |
| Gestational age | -MOBA | 0.0002 | 0.0001 | 0.0004 | 0.0000 | 0.040 | 0.074 | 0.005 | 0.02 | 0.33 |
| | -Public | 0.0002 | 0.0002 | 0.0005 | -0.0001 | 0.046 | 0.109 | -0.018 | 0.16 | 0.62 |

| | | | | | | | | | | |
|---|---|---|---|---|---|---|---|---|---|---|
| | -UKB | 0.0002 | 0.0001 | 0.0004 | 0.0001 | 0.044 | 0.076 | 0.012 | 0.01 | 0.33 |
| Birth weight | -ALSPAC | 0.0000 | 0.0000 | 0.0001 | 0.0000 | 0.006 | 0.019 | -0.006 | 0.31 | 0.73 |
| | -BIB-SA | 0.0000 | 0.0000 | 0.0001 | 0.0000 | 0.006 | 0.018 | -0.007 | 0.37 | 0.74 |
| | -BIB-WE | 0.0000 | 0.0000 | 0.0001 | 0.0000 | 0.006 | 0.018 | -0.007 | 0.35 | 0.74 |
| | -MOBA | 0.0000 | 0.0000 | 0.0001 | 0.0000 | 0.004 | 0.018 | -0.009 | 0.54 | 0.86 |
| | -UKB | 0.0001 | 0.0000 | 0.0001 | 0.0000 | 0.013 | 0.041 | -0.015 | 0.36 | 0.74 |

**Table S7*: Results of the genetic colocalization analysis between Lp(a) levels and adverse pregnancy APPOs within the LPA locus.*** For the definitions of the hypothesis underlying the posterior probabilities please refer to Section 2.6.4. NICU – neonatal intensive care unit.

| APPO | PP.H0 | PP.H1 | PP.H2 | PP.H3 | PP.H4 |
|---|---|---|---|---|---|
| Hypertensive disorders of pregnancy | 0.00 | 0.94 | 0.00 | 0.04 | 0.02 |
| Gestational hypertension | 0.00 | 0.89 | 0.00 | 0.07 | 0.04 |
| Preeclampsia | 0.00 | 0.81 | 0.00 | 0.17 | 0.01 |
| Gestational diabetes | 0.00 | 0.93 | 0.00 | 0.06 | 0.01 |
| Miscarriage | 0.00 | 0.93 | 0.00 | 0.07 | 0.01 |
| Recurrent miscarriage | 0.00 | 0.84 | 0.00 | 0.14 | 0.02 |
| Stillbirth | 0.00 | 0.88 | 0.00 | 0.08 | 0.03 |
| Induction of labour | 0.00 | 0.75 | 0.00 | 0.20 | 0.04 |
| Pre-labour rupture of membrane | 0.00 | 0.93 | 0.00 | 0.06 | 0.01 |
| Caesarian section | 0.00 | 0.88 | 0.00 | 0.10 | 0.02 |
| Pre-term birth | 0.00 | 0.90 | 0.00 | 0.06 | 0.04 |
| Small for gestational age | 0.00 | 0.75 | 0.00 | 0.24 | 0.01 |
| Large for gestational age | 0.00 | 0.89 | 0.00 | 0.10 | 0.01 |
| Low Apgar score at 1 minute | 0.00 | 0.82 | 0.00 | 0.07 | 0.11 |

| | | | | | |
|---|---|---|---|---|---|
| Low Apgar score at 5 minutes | 0.00 | 0.75 | 0.00 | 0.22 | 0.03 |
| Postpartum haemorrhage | 0.00 | 0.89 | 0.00 | 0.09 | 0.01 |
| NICU admission | 0.00 | 0.90 | 0.00 | 0.09 | 0.02 |

**Table S8: Mrlap estimates for the estimated effect of Lp(a) lowering on adverse pregnancy and perinatal outcomes.** All estimates refer to the effect of lowering Lp(a) by 1nmol/L in log odds ratio or mean difference scale. Note that this analysis considered the entire genome and focused on detecting bias from sample overlap.

| outcome | nsnp | Observed beta | Observed SE | Observed P-value | Corrected beta | Corrected SE | Corrected P-value | Test difference | P-value test difference |
|---|---|---|---|---|---|---|---|---|---|
| Miscarriage | 10 | -0.0135 | 0.0075 | 0.07 | -0.0135 | 0.0109 | 0.21 | -0.0028 | 1.00 |
| Recurrent miscarriage | 10 | -0.0038 | 0.0094 | 0.69 | -0.0038 | 0.0093 | 0.69 | -0.0002 | 1.00 |
| Stillbirth | 10 | -0.0005 | 0.0117 | 0.97 | -0.0005 | 0.0117 | 0.97 | 0.0030 | 1.00 |
| Hypertensive disorders of pregnancy | 9 | -0.0030 | 0.0059 | 0.61 | -0.0030 | 0.0057 | 0.60 | -0.0030 | 1.00 |
| Gestational hypertension | 9 | -0.0020 | 0.0059 | 0.74 | -0.0020 | 0.0061 | 0.74 | 0.0012 | 1.00 |
| Preeclampsia | 10 | -0.0155 | 0.0112 | 0.17 | -0.0155 | 0.0115 | 0.18 | -0.0101 | 0.99 |
| Gestational diabetes | 9 | -0.0078 | 0.0052 | 0.13 | -0.0078 | 0.0052 | 0.13 | -0.0543 | 0.96 |
| Induction of labour | 10 | -0.0332 | 0.0146 | 0.02 | -0.0332 | 0.0145 | 0.02 | -0.0286 | 0.98 |
| Pre-labour rupture of membrane | 10 | -0.0128 | 0.0125 | 0.31 | -0.0128 | 0.0124 | 0.30 | -0.0079 | 0.99 |
| Caesarian section | 9 | 0.0073 | 0.0093 | 0.43 | 0.0074 | 0.0702 | 0.92 | 0.0001 | 1.00 |
| Birth weight | 10 | 0.0055 | 0.0105 | 0.60 | 0.0055 | 0.0105 | 0.60 | 0.0232 | 0.98 |
| Gestational age | 10 | 0.0312 | 0.0125 | 0.01 | 0.0312 | 0.0135 | 0.02 | 0.0098 | 0.99 |

| | | | | | | | | | |
|---|---|---|---|---|---|---|---|---|---|
| Spontaneous pre-term birth | 10 | -0.0053 | 0.0107 | 0.62 | -0.0054 | 0.0107 | 0.62 | -0.0108 | 0.99 |
| Small for gestational age | 9 | -0.0188 | 0.0145 | 0.20 | -0.0188 | 0.0146 | 0.20 | -0.1087 | 0.91 |
| Large for gestational age | 10 | 0.0148 | 0.0185 | 0.42 | 0.0149 | 0.0184 | 0.42 | 0.0059 | 1.00 |
| Low Apgar score at 1 minute | 10 | 0.0035 | 0.0178 | 0.85 | 0.0035 | 0.0197 | 0.86 | -0.0007 | 1.00 |
| Low Apgar score at 5 minutes | 10 | -0.0297 | 0.0164 | 0.07 | -0.0298 | 0.7804 | 0.97 | -0.0001 | 1.00 |
| NICU admission | 10 | -0.0299 | 0.0161 | 0.06 | -0.0300 | 0.3196 | 0.93 | -0.0002 | 1.00 |
| Congenital malformations (any) | 10 | 0.0118 | 0.0165 | 0.47 | 0.0118 | 0.0345 | 0.73 | 0.0003 | 1.00 |

**Table S9: Positive control MR estimates for the estimated effect of Lp(a) lowering on coronary artery disease.** $^{1}$ denotes MR estimates reflecting the effect of lowering Lp(a) by 1 nmol/L, $^{-210}$ denotes MR estimates reflecting the effect of lowering Lp(a) by 210 nmol/L. OR, odds ratio (binary outcome); SE, standard error of beta/log odds estimate; UCI, upper 95% confidence interval; LCI, lower 95% confidence interval.

| Outcome | nsnp | F | $r^2$ | $OR^{(-1)}$ | $SE^{(-1)}$ | $UCI^{(-1)}$ | $LCI^{(-1)}$ | $OR^{(-210)}$ | $UCI^{(-210)}$ | $LCI^{(-210)}$ | P-value |
|---|---|---|---|---|---|---|---|---|---|---|---|
| CAD | 13 | 18961.8 | 0.64 | 0.9971 | 0.0004 | 0.9979 | 0.9962 | 0.548 | 0.653 | 0.459 | 2.13E-11 |

##### Table S10: Comparison of BMI and smoking across quintiles of the Lp(a) genetic score.

| GS quintile | Total N | BMI mean | BMI SD | Missing BMI | Never smoked | Used to smoke | Current smoker | Missing smoking status |
|---|---|---|---|---|---|---|---|---|
| 1 | 39771 | 27.0194375 | 5.06469628 | 4690 | 20135 | 11735 | 3181 | 4720 |
| 2 | 39771 | 27.0470974 | 5.05351605 | 4710 | 20380 | 11591 | 3076 | 4724 |
| 3 | 39771 | 27.0159014 | 5.05537657 | 4820 | 20089 | 11719 | 3094 | 4869 |
| 4 | 39771 | 27.0514716 | 5.08934916 | 4657 | 20283 | 11694 | 3133 | 4661 |
| 5 | 39770 | 27.0039253 | 5.09036053 | 4779 | 20339 | 11512 | 3136 | 4783 |

**Table S11:** ***Reported reason for missing Lp(a) measurement across each of the Lp(a) genetic score quintile.*** The reportable range for the UKB Lp(a) assay was 3.8 - 189 nmol/L.

| Genetic score quintile | Lp(a) not measured | Lp(a) above assay limit | Lp(a) below assay limit | Lp(a) within range |
|---|---|---|---|---|
| 1 | 7336 | 548 | 3527 | 28360 |
| 2 | 7257 | 583 | 5932 | 25999 |
| 3 | 7443 | 747 | 3428 | 28153 |
| 4 | 7197 | 1511 | 1635 | 29428 |
| 5 | 7437 | 9798 | 547 | 21988 |

**Table S12:** ***Case numbers for each APPO across the Lp(a) genetic score quintiles.*** Each quintile had 35097 individuals. Counts under 5 are presented as "<5" to abide by UKB regulations.

| Outcome | Q1 | Q2 | Q3 | Q4 | Q5 |
|---|---|---|---|---|---|
| Any thrombotic event (venous + arterial) | 6 | <5 | 5 | <5 | 6 |
| Venous thrombotic events | 6 | <5 | 5 | <5 | 6 |
| Arterial thrombotic events | 0 | 0 | 0 | 0 | 0 |
| Hypertensive disorders of pregnancy | 1664 | 1724 | 1689 | 1775 | 2206 |
| Gestational hypertension | 127 | 146 | 125 | 129 | 143 |
| Preeclampsia | 74 | 106 | 69 | 88 | 92 |
| Gestational diabetes | 68 | 54 | 67 | 52 | 59 |
| Miscarriage | 164 | 154 | 158 | 151 | 161 |
| Recurrent miscarriage | 8613 | 8475 | 8474 | 8568 | 8540 |
| Stillbirth | 757 | 722 | 751 | 769 | 748 |
| Induction of labour | 989 | 967 | 1002 | 1005 | 940 |
| Caesarian section | 346 | 364 | 301 | 338 | 375 |
| Spontaneous pre-term birth | 397 | 400 | 421 | 397 | 371 |

| | | | | | |
|---|---|---|---|---|---|
| Small for gestational age | 42 | 49 | 45 | 42 | 44 |
| Large for gestational age | 87 | 82 | 74 | 77 | 75 |

**Table S13: Estimated association of each Lp(a) genetic score quintile with each of the APPOs, relative to the highest quintile.** OR, odds ratio (binary outcome); beta, mean difference (continuous outcome); SE, standard error of beta/log odds estimate; UCI, upper 95% confidence interval; LCI, lower 95% confidence interval; FDR, False discovery rate.

| Outcome | Quintile being compared to Q5 | OR/beta | SE | UCI | LCI | P-value | FDR P-value |
|---|---|---|---|---|---|---|---|
| Hypertensive disorders of pregnancy | Q1 | 0.799 | 0.123 | 1.017 | 0.628 | 0.07 | 0.48 |
| | Q2 | 0.934 | 0.118 | 1.177 | 0.741 | 0.56 | 0.96 |
| | Q3 | 0.817 | 0.122 | 1.039 | 0.644 | 0.10 | 0.52 |
| | Q4 | 0.810 | 0.122 | 1.029 | 0.637 | 0.08 | 0.48 |
| Gestational hypertension | Q1 | 0.730 | 0.157 | 0.993 | 0.537 | 0.04 | 0.44 |
| | Q2 | 1.028 | 0.143 | 1.361 | 0.777 | 0.85 | 0.97 |
| | Q3 | 0.747 | 0.155 | 1.013 | 0.551 | 0.06 | 0.48 |
| | Q4 | 0.871 | 0.149 | 1.167 | 0.651 | 0.35 | 0.83 |
| Preeclampsia | Q1 | 1.058 | 0.180 | 1.506 | 0.743 | 0.75 | 0.97 |
| | Q2 | 0.884 | 0.188 | 1.279 | 0.611 | 0.51 | 0.96 |
| | Q3 | 0.991 | 0.182 | 1.417 | 0.693 | 0.96 | 0.99 |
| | Q4 | 0.805 | 0.193 | 1.175 | 0.552 | 0.26 | 0.68 |
| Gestational diabetes | Q1 | 0.969 | 0.111 | 1.205 | 0.780 | 0.78 | 0.97 |
| | Q2 | 0.937 | 0.112 | 1.167 | 0.753 | 0.56 | 0.96 |
| | Q3 | 0.970 | 0.111 | 1.206 | 0.781 | 0.78 | 0.97 |
| | Q4 | 0.938 | 0.112 | 1.168 | 0.754 | 0.57 | 0.96 |

| | | | | | | | |
|---|---|---|---|---|---|---|---|
| Miscarriage | Q1 | 0.992 | 0.018 | 1.028 | 0.957 | 0.66 | 0.97 |
| | Q2 | 0.975 | 0.018 | 1.010 | 0.941 | 0.16 | 0.57 |
| | Q3 | 0.973 | 0.018 | 1.008 | 0.939 | 0.13 | 0.54 |
| | Q4 | 0.988 | 0.018 | 1.024 | 0.954 | 0.52 | 0.96 |
| Recurrent miscarriage | Q1 | 1.033 | 0.053 | 1.146 | 0.931 | 0.54 | 0.96 |
| | Q2 | 0.973 | 0.054 | 1.081 | 0.876 | 0.61 | 0.96 |
| | Q3 | 1.008 | 0.053 | 1.118 | 0.908 | 0.88 | 0.97 |
| | Q4 | 1.035 | 0.053 | 1.148 | 0.933 | 0.52 | 0.96 |
| Stillbirth | Q1 | 1.000 | 0.047 | 1.095 | 0.912 | 0.99 | 0.99 |
| | Q2 | 0.989 | 0.047 | 1.084 | 0.902 | 0.81 | 0.97 |
| | Q3 | 1.024 | 0.046 | 1.122 | 0.935 | 0.61 | 0.96 |
| | Q4 | 1.002 | 0.047 | 1.097 | 0.914 | 0.97 | 0.99 |
| Induction of labour | Q1 | 0.859 | 0.087 | 1.019 | 0.724 | 0.08 | 0.48 |
| | Q2 | 0.857 | 0.086 | 1.015 | 0.724 | 0.07 | 0.48 |
| | Q3 | 0.711 | 0.089 | 0.846 | 0.598 | 0.00 | 0.00 |
| | Q4 | 0.874 | 0.087 | 1.037 | 0.736 | 0.12 | 0.54 |
| Caesarian section | Q1 | 1.105 | 0.086 | 1.307 | 0.934 | 0.24 | 0.68 |
| | Q2 | 1.049 | 0.085 | 1.240 | 0.887 | 0.58 | 0.96 |
| | Q3 | 1.130 | 0.084 | 1.334 | 0.958 | 0.15 | 0.57 |
| | Q4 | 1.129 | 0.086 | 1.336 | 0.955 | 0.16 | 0.57 |

| | | | | | | | |
|---|---|---|---|---|---|---|---|
| Pre-term birth | Q1 | 1.047 | 0.226 | 1.629 | 0.672 | 0.84 | 0.97 |
| | Q2 | 1.181 | 0.219 | 1.814 | 0.768 | 0.45 | 0.96 |
| | Q3 | 1.039 | 0.222 | 1.605 | 0.673 | 0.86 | 0.97 |
| | Q4 | 1.045 | 0.223 | 1.617 | 0.675 | 0.84 | 0.97 |
| Small for gestational age | Q1 | 1.221 | 0.166 | 1.690 | 0.882 | 0.23 | 0.68 |
| | Q2 | 1.050 | 0.168 | 1.460 | 0.755 | 0.77 | 0.97 |
| | Q3 | 1.044 | 0.169 | 1.453 | 0.749 | 0.80 | 0.97 |
| | Q4 | 1.082 | 0.170 | 1.508 | 0.776 | 0.64 | 0.97 |
| Large for gestational age | Q1 | 1.008 | 0.112 | 1.255 | 0.809 | 0.94 | 0.99 |
| | Q2 | 0.947 | 0.112 | 1.179 | 0.760 | 0.62 | 0.96 |
| | Q3 | 0.986 | 0.111 | 1.227 | 0.793 | 0.90 | 0.97 |
| | Q4 | 1.080 | 0.111 | 1.342 | 0.869 | 0.49 | 0.96 |
| Gestational age | Q1 | 0.093 | 0.067 | 0.225 | -0.039 | 0.17 | 0.57 |
| | Q2 | 0.141 | 0.067 | 0.272 | 0.011 | 0.03 | 0.38 |
| | Q3 | 0.076 | 0.067 | 0.207 | -0.055 | 0.25 | 0.68 |
| | Q4 | 0.078 | 0.067 | 0.210 | -0.054 | 0.25 | 0.68 |
| Birth weight | Q1 | 0.012 | 0.008 | 0.027 | -0.003 | 0.13 | 0.54 |
| | Q2 | 0.002 | 0.008 | 0.017 | -0.013 | 0.83 | 0.97 |
| | Q3 | 0.002 | 0.008 | 0.017 | -0.013 | 0.81 | 0.97 |
| | Q4 | 0.009 | 0.008 | 0.024 | -0.006 | 0.25 | 0.68 |

| | | | | | | | |
|---|---|---|---|---|---|---|---|
| Venous thrombotic event | Q1 | 1.118 | 0.559 | 3.348 | 0.374 | 0.84 | 0.97 |
| | Q2 | 0.480 | 0.711 | 1.933 | 0.119 | 0.30 | 0.73 |
| | Q3 | 0.940 | 0.581 | 2.934 | 0.301 | 0.92 | 0.97 |
| | Q4 | 0.661 | 0.647 | 2.352 | 0.186 | 0.52 | 0.96 |
| Coronary artery disease | Q1 | 0.734 | 0.034 | 0.784 | 0.687 | 0.00 | 0.00 |
| | Q2 | 0.776 | 0.033 | 0.827 | 0.727 | 0.00 | 0.00 |
| | Q3 | 0.754 | 0.033 | 0.804 | 0.706 | 0.00 | 0.00 |
| | Q4 | 0.797 | 0.033 | 0.850 | 0.748 | 0.00 | 0.00 |

**Table S14: MAF (minor allele frequency) observed in the UK Biobank for LPA LoF variants.**

| Variant | MAF |
|---|---|
| rs41267811_C | 0.000128 |
| rs143431368_C | 0.002469 |
| rs200099994_T | 0.000194 |
| rs41267813_A | 0.001125 |
| rs41272114_T | 0.038159 |
| rs41259144_T | 0.010628 |
| rs559580002_T | 0.000028 |

**Table S15: *Reported reason for missing Lp(a) measurement across each of the Lp(a) genetic score quintile.*** The reportable range for the UKB Lp(a) assay was 3.8 - 189 nmol/L.

| LoF status | Not measured | Levels above assay limits | Levels below assay limits | Levels within assay range |
|---|---|---|---|---|
| Heterozygote/wild-type | 36445 | 13102 | 14700 | 133353 |
| Homozygote | 70 | 0 | 286 | 6 |

Table S16: Number of APPO cases per LoF status. Counts under 5 are presented as ‘<5’ to abide by UKB regulations.

| Outcome | Knockouts (total N = 362) | Rest of sample (total N = 197,600) |
|---|---|---|
| Any thrombotic event | 0 | 22 |
| Venous thrombotic event | 0 | 22 |
| Atherosclerotic thrombotic event | 0 | 0 |
| Coronary artery disease | 9 | 9017 |
| Hypertensive disorders of pregnancy | <5 | 667 |
| Gestational hypertension | 0 | 428 |
| Preeclampsia | <5 | 298 |
| Gestational diabetes | <5 | 783 |
| Miscarriage | 78 | 42410 |
| Recurrent miscarriage | 8 | 3725 |
| Stillbirth | 8 | 4877 |
| Induction of labour | <5 | 1719 |
| Caesarian section | <5 | 1976 |
| Spontaneous pre-term birth | 0 | 222 |
| Small for gestational age | <5 | 394 |
| Large for gestational age | <5 | 972 |

**Table S17: Estimated association of LPA knockout with each of the APPOs, relative to the rest of the sample (heterozygotes and wild-types for LPA LoF variants).** CI – 95% confidence interval.

| **Outcome** | **OR/beta** | **se** | **Lower CI** | **Upper CI** | ***P* value** | **FDR** |
|---|---|---|---|---|---|---|
| Hypertensive disorders of pregnancy | 0.8086 | 1.0031 | 0.1132 | 5.7750 | 0.83 | 0.99 |
| Preeclampsia | 1.7600 | 1.0043 | 0.2458 | 12.6011 | 0.57 | 0.99 |
| Gestational diabetes | 1.4417 | 0.7106 | 0.3581 | 5.8047 | 0.61 | 0.99 |
| Miscarriage | 1.0593 | 0.1351 | 0.8128 | 1.3805 | 0.67 | 0.99 |
| Recurrent miscarriage | 1.2236 | 0.3625 | 0.6012 | 2.4903 | 0.58 | 0.99 |
| Stillbirth | 0.9434 | 0.3616 | 0.4644 | 1.9163 | 0.87 | 0.99 |
| Induction of labour | 0.2543 | 1.0415 | 0.0330 | 1.9582 | 0.19 | 0.99 |
| Caesarian section | 1.2190 | 0.6042 | 0.3730 | 3.9840 | 0.74 | 0.99 |
| Small for gestational age | 1.2634 | 1.0529 | 0.1604 | 9.9494 | 0.82 | 0.99 |
| Large for gestational age | 1.0909 | 0.7828 | 0.2352 | 5.0598 | 0.91 | 0.99 |
| Gestational age | -0.0590 | 0.4809 | -1.0017 | 0.8836 | 0.90 | 0.99 |
| Birth weight | -0.0813 | 0.0571 | -0.1932 | 0.0305 | 0.15 | 0.99 |
| Coronary artery disease | 0.5280 | 0.3387 | 0.2718 | 1.0256 | 0.06 | 0.99 |

#### Supplementary Figures

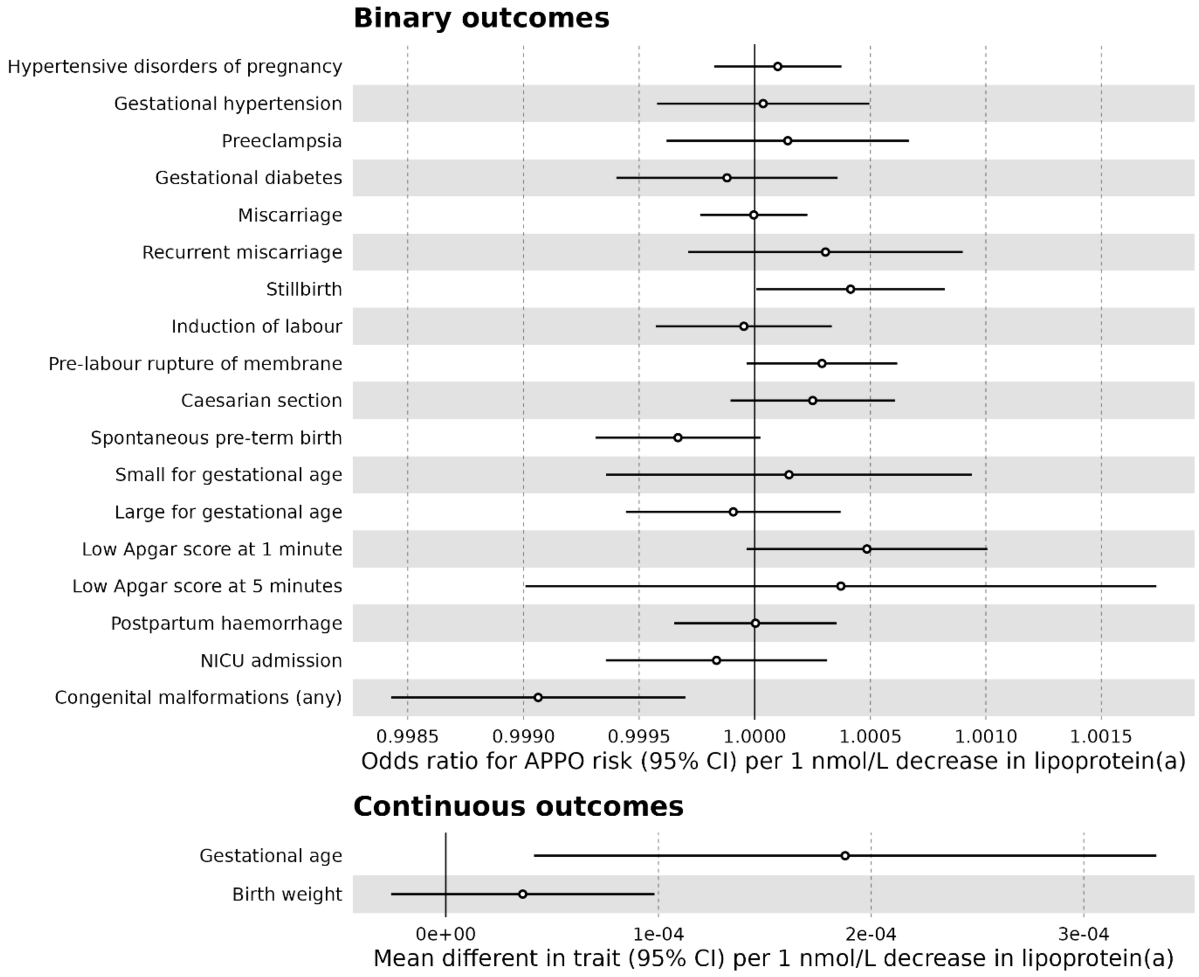


**Figure S1: Mendelian randomization estimates for the estimated effect of Lp(a) lowering on adverse pregnancy and perinatal outcomes for a genetically-predicted decrease of Lp(a) by 1 nmol/L.** Results are expressed as odds ratio (binary outcomes) or mean difference in trait (continuous outcomes) for every 1 nmol/L decrease in Lp(a) resulting from genetically-instrumented downregulation of the drug target gene (LPA). Point estimates and 95% confidence intervals are represented by circles and bars, respectively. Open circles indicate FDR-corrected P > 0.05. Gestational age is in weeks and birth weight is in Z units.

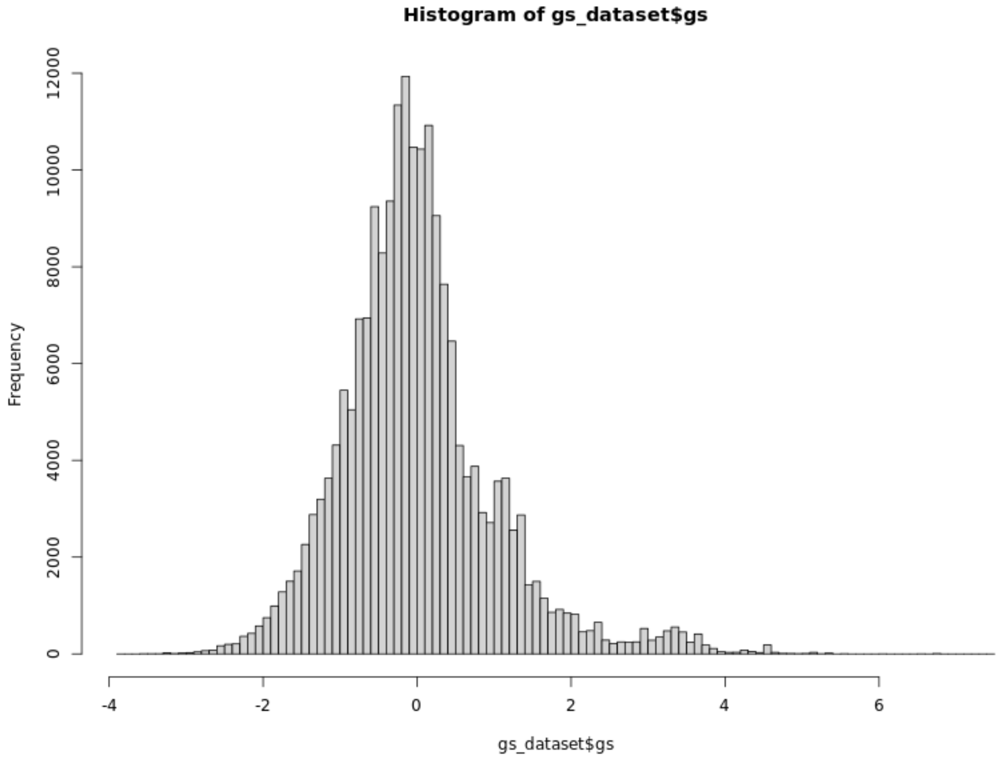


**Figure S2: Distribution of the Lp(a) genetic score in the MR-PREG UKB sample, after scaling it so mean is 0 and standard deviation is 1.**

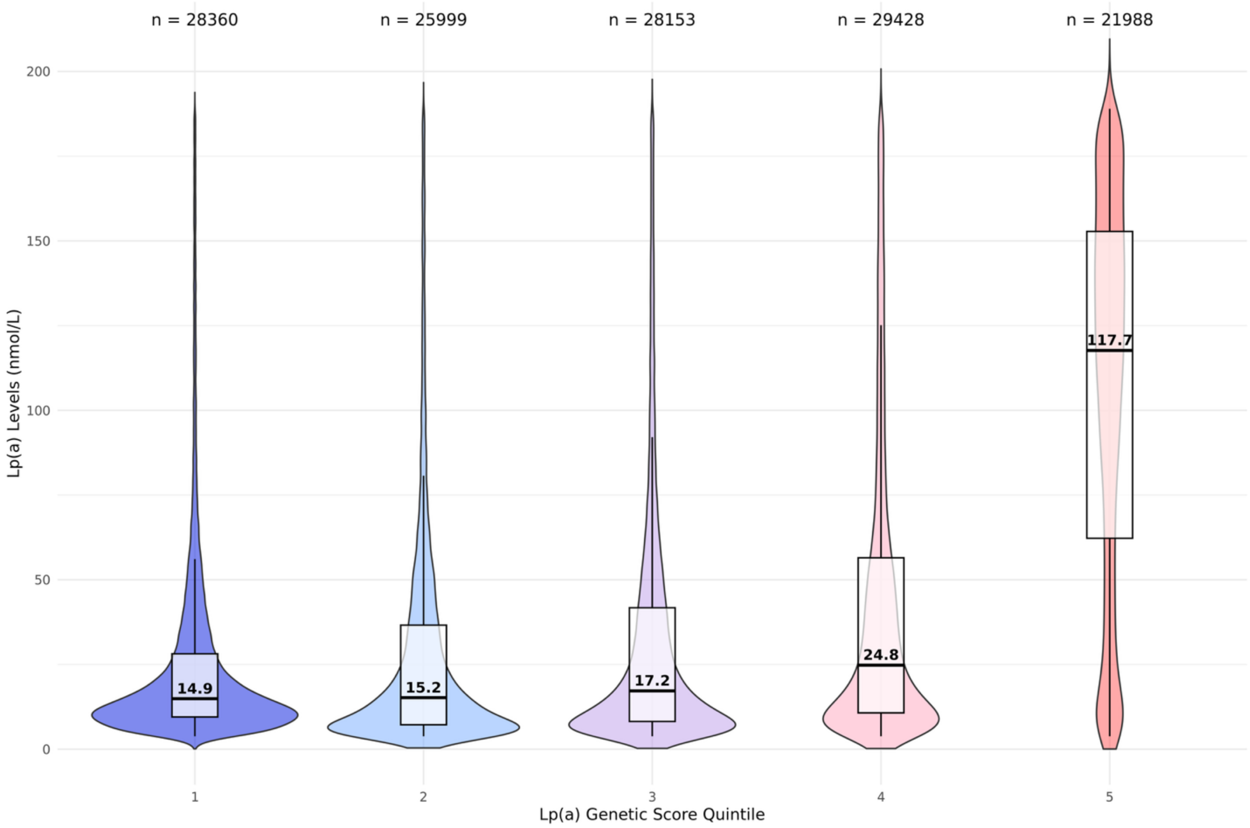


**Figure S3: Violin and box plot of measured Lp(a) levels across the Lp(a) genetic score quintiles.**

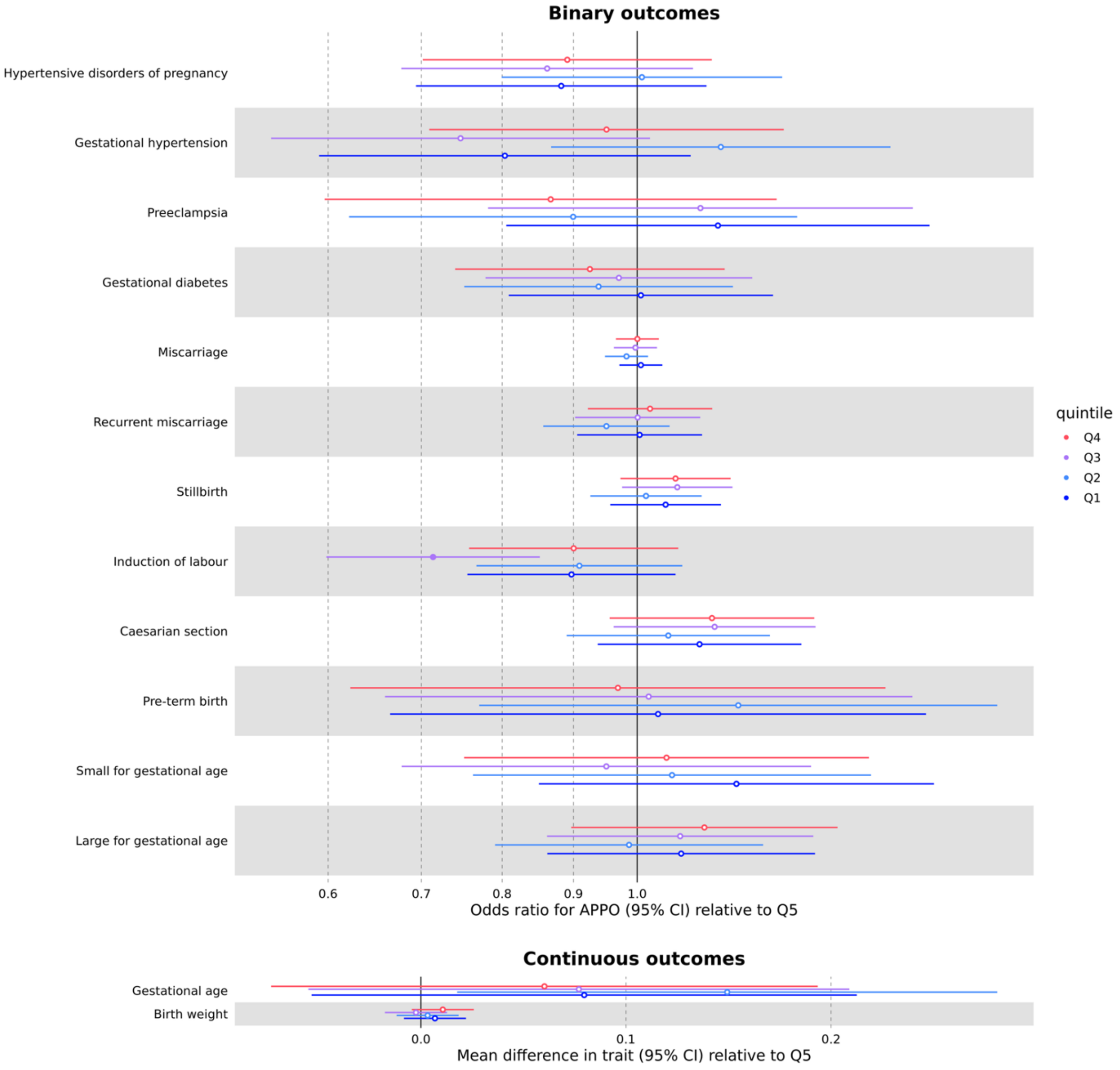


**Figure S4: Estimated association of each Lp(a) genetic score quintile with each of the APPOs, relative to the highest quintile.** Models were adjusted for the first 40 genetic principal components and genotyping batch. Point estimates and 95% confidence intervals are represented by circles and bars, respectively. Open circles indicate FDR-corrected P > 0.05. Gestational age is in weeks and birth weight is in Z units. Thrombotic events are excluded from this plot due to the extreme estimates caused by low case numbers (see Table 7).

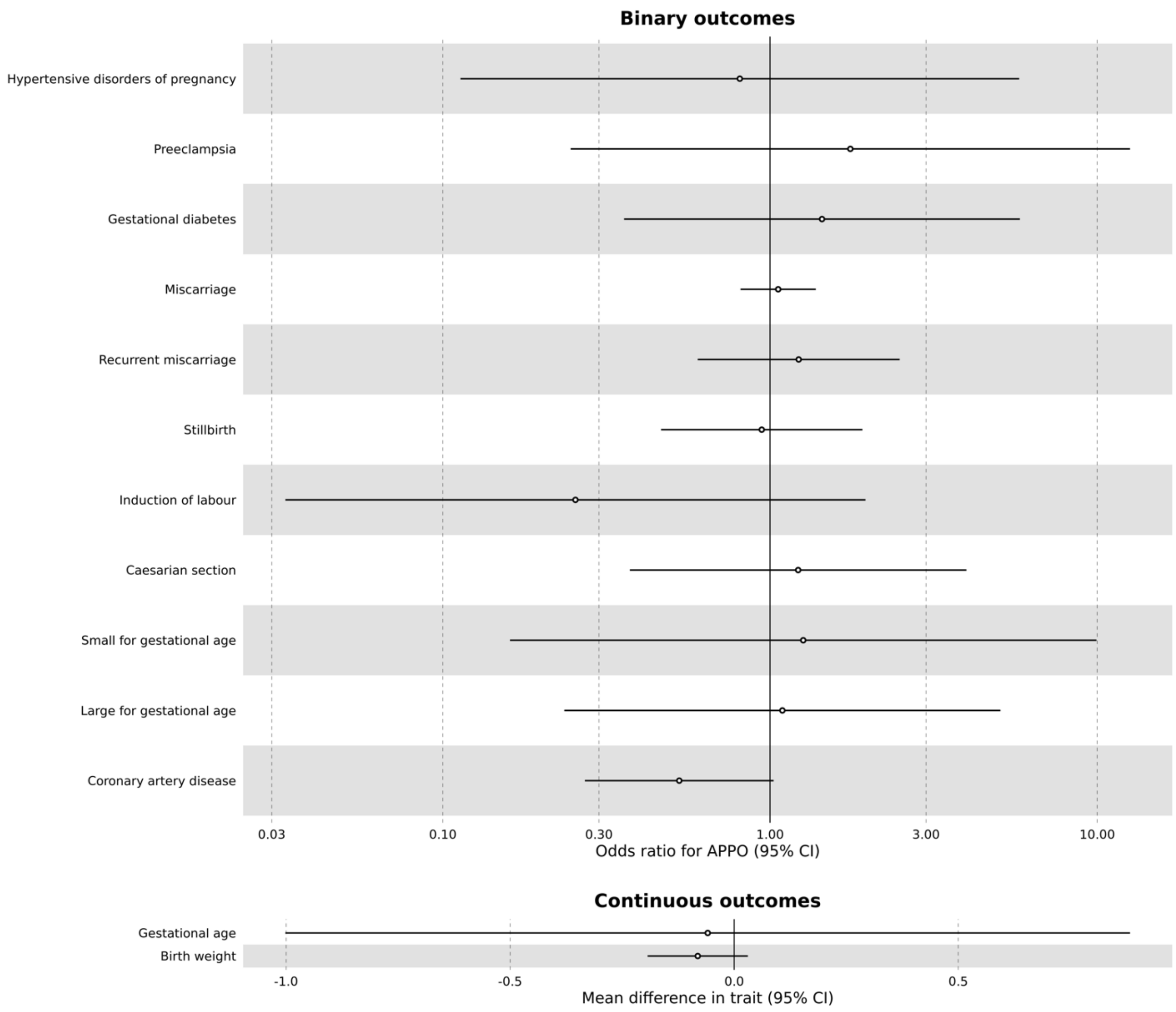


**Figure S5: Estimated association of LPA knockout with each of the APPOs, relative to the rest of the sample (heterozygotes and wild-types for LPA LoF variants).** Models were adjusted for the first 40 genetic principal components and genotyping batch. Point estimates and 95% confidence intervals are represented by circles and bars, respectively. Open circles indicate FDR-corrected P > 0.05. Gestational age is in weeks and birth weight is in Z units. Thrombotic events were excluded from this analysis due to having no cases in the LPA knockout group. Coronary artery disease in the general population is added to the figure as a positive control outcome.

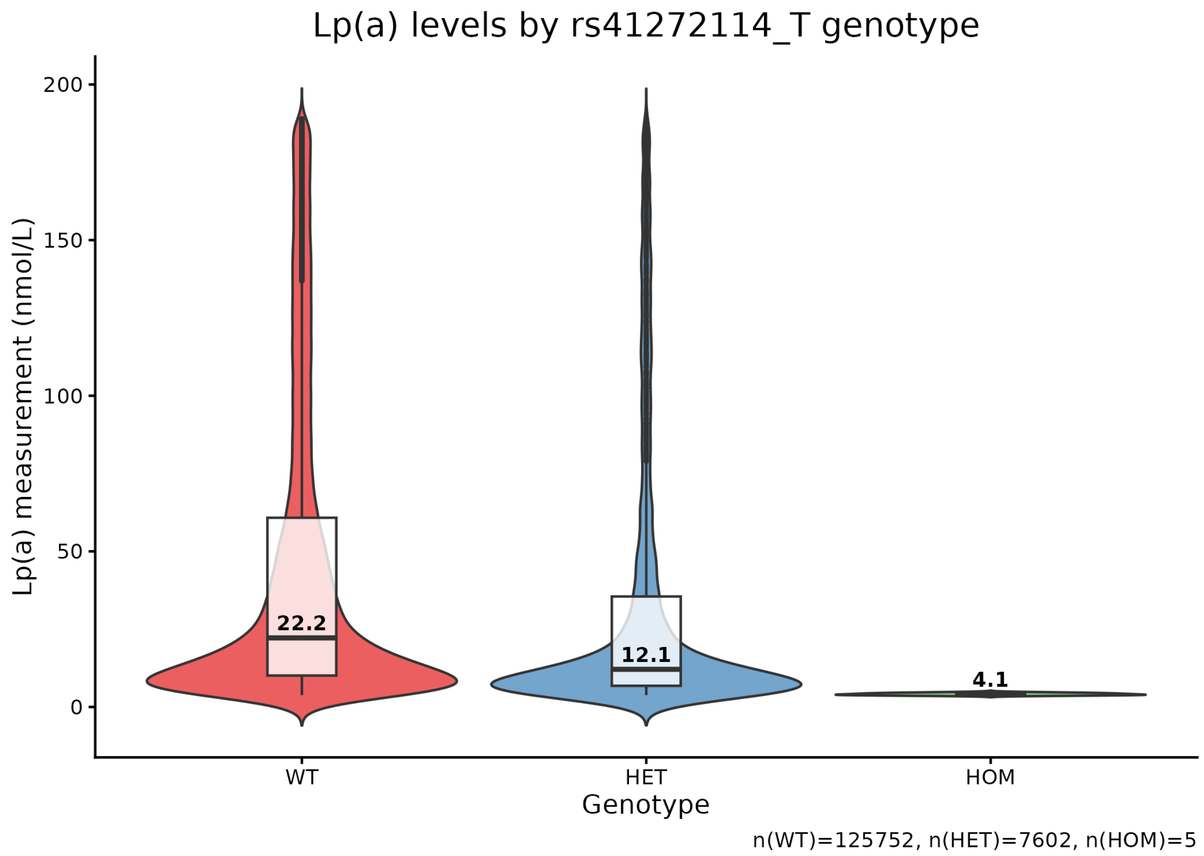


**Figure S6: Comparison of measured Lp(a) levels according to rs41272114 genotype.** WT, wild-type; HET, heterozygote; HOM – homozygote/knockout.
